# Midlife Health in Britain and the US: A comparison of Two Nationally Representative Cohorts

**DOI:** 10.1101/2023.12.21.23300366

**Authors:** Charis Bridger Staatz, Iliya Gutin, Andrea Tilstra, Laura Gimeno, Bettina Moltrecht, Dario Moreno-Agostino, Vanessa Moulton, Martina K. Narayanan, Jennifer B. Dowd, Lauren Gaydosh, George B. Ploubidis

## Abstract

**Background:** Older adults in the United States (US) have worse health and wider socioeconomic inequalities in health compared to Britain. Less is known about how health in the two countries compares in midlife, a time of emerging health decline, including inequalities in health.

**Methods:** We compare measures of smoking status, alcohol consumption, obesity, self-rated health, cholesterol, blood pressure, and glycated haemoglobin using population-weighted modified Poisson regression in the 1970 British Cohort Study (BCS70) in Britain (N= 9,665) and the National Longitudinal Study of Adolescent to Adult Health (Add Health) in the US (N=12,297), when cohort members were aged 34-46 and 33-43, respectively. We test whether associations vary by early- and mid-life socioeconomic position.

**Findings:** US adults had higher levels of obesity, high blood pressure and high cholesterol. Prevalence of poor self-rated health, heavy drinking, and smoking was worse in Britain. We found smaller socioeconomic inequalities in midlife health in Britain compared to the US. For some outcomes (e.g., smoking), the most socioeconomically advantaged group in the US was healthier than the equivalent group in Britain. For other outcomes (hypertension and cholesterol), the most advantaged US group fared equal to or worse than the most disadvantaged groups in Britain.

**Interpretation:** US adults have worse cardiometabolic health than British counterparts, even in early midlife. The smaller socioeconomic inequalities and better overall health in Britain may reflect differences in access to health care, welfare systems, or other environmental risk factors.

**Funding:** ESRC, UKRI, MRC, NIH, European Research Council, Leverhulme Trust

**Research in context:** *Evidence before this study:* This study considered a range of seminal evidence published in academic journals, focusing on international comparisons of health, of which the majority has been conducted in older age cohorts (adults over the age of 50) in Britain and the US. We focused our search on cross-country comparisons and international surveys of ageing, such as the Health and Retirement Survey in the US, and the English Longitudinal Study of Ageing in Britain. We limited our search to English language publications and included studies that considered both overall differences in health, and differences in socioeconomic inequalities in health. The majority of considered studies found older adults in the US to have worse health than in Britain, and with greater evidence of inequalities for older adults in the US. However, older adults in Britain were more likely to exhibit worse health behaviours than those in the US.

*Added value of this study:* This study adds value by investigating health in early midlife (30s and 40s), a period less researched compared to older age. Midlife is an important time in the life course where early signs of decline can be observed and when there is still an opportunity to promote healthy aging. The importance of midlife is consistent with the need to understand healthy ageing as a life-long process. This study uses biomarkers as objective measures of cardiometabolic health and involved retrospective harmonisation of cohorts in Britain and the US, helping lay the groundwork for efforts to harmonise cohorts at younger ages and facilitate comparative work.

*Implications of all the available evidence:* We find that health in US adults is worse than their peers in Britain at even earlier ages (30s-40s years of age) than previously documented, especially for cardiometabolic measures. While associations of childhood socioeconomic status and later health were found in both Britain and the US, adult socioeconomic measures largely accounted for these associations. This finding is consistent with previous work and underscores the persistence of socioeconomic position across the life course, with sustained impacts on health. Policies aimed at improving health must consider this link between early and later life socioeconomic circumstances. We also find wider socioeconomic inequalities in health outcomes in the US than Britain. For some outcomes the most advantaged groups in the US have similar or worse health than the most disadvantaged groups in Britain. These findings, along with previously published evidence, have implications for policy and practice, as they suggest sociopolitical differences between the two countries that may drive different health profiles. Systematic differences between Britain and the US in terms of health care and welfare provisions may drive both worse health, and wider inequalities in the US.

## Background

International comparisons document worse health in the United States (US) compared to England (1–5). Older US adults are less healthy based on measures of self-reported diabetes, hypertension, heart disease, myocardial infarction, stroke, lung disease, and cancer (2). They also have a higher average body mass index and prevalence of extreme obesity (4). However, older adults in England exhibit worse health behaviours, including the co-occurrence of smoking, alcohol consumption, and low physical activity, and were less likely to present with no behavioural risk factors (1).

Previous comparisons of health in the US and England have primarily focused on ages 50 (late midlife) or 60 and over (old age) based on harmonised international surveys of ageing (6). Midlife (ages 30-60), and particularly younger midlife (ages 30-40), is often overlooked in life course research on health (7). Yet, there is growing recognition of midlife as an important period that sets the stage for later life health and aging, marking the start of physical and functional decline (8). In contrast to US-England health differences at older ages, the limited evidence at younger ages is more equivocal. One comparison finds similar patterns of worse cardiometabolic health but better health behaviours in the US compared to England at ages 35-54 (5). Conversely, a more recent study examining individuals born between 1965-80 documents higher prevalence of hypertension and dyslipidaemia among English adults and comparable prevalence of smoking and diabetes risk (9, 10).

Moreover, these midlife declines in health likely exhibit social gradients. Studies comparing England and the US document wealth, income, and education inequalities across multiple chronic conditions for adults in their 50s and 60s (2, 3), with some evidence of similar gradients at midlife (5, 11). In both countries, behavioral risk factors (obesity, exercise, and smoking) explain some of these gradients, but a sizeable proportion remains unaccounted for – indicative of the multiple mechanisms through which social disadvantages operate (3, 5).

Many of these health inequalities originate even earlier in the life course, as multiple studies of British and US adults find strong associations between early life socioeconomic position (SEP) and adult health (12, 13).

Critically, international comparisons provide the opportunity to identify contextual drivers of population health, (14–16) with prior work finding smaller health inequalities in countries with higher national incomes, social transfers and health care expenditure and quality of health care and relevant policy (14). Observed differences between the US and England have been attributed to the cost of healthcare (2, 5), which is free at the point of access in the UK (England, Scotland, Wales and Northern Ireland), differences in income benefit systems (2, 3) and the quality of local environments and neighbourhoods (17). These contextual determinants of health inequalities likely vary across the life course as the level of welfare provisions differs between countries and life stages (e.g., retirement benefits and childhood welfare).

Thus, we build on previous work and compare behavioural risk factors and biomarkers of health in early midlife from two nationally representative cohorts, the National Longitudinal Study of Adolescent to Adult Health (Add Health) in the US, and the 1970 British Cohort study (BCS70) in Britain (England, Wales, and Scotland). We consider the moderating role of early life SEP and current SEP. Drawing on past work, we hypothesise that the health of the US cohort will generally be worse than their British peers, and that US SEP inequalities will be larger.

## Methods

### Data Sets

BCS70 is an ongoing nationally representative birth cohort of ∼17,000 individuals born in 1970 in Britain (18). Cohort members have been followed up ten times since birth. The tenth sweep, in 2016, collected multiple biomedical measures, including blood samples. The current analysis uses data from sweeps eight to ten, when cohort members were ages 34 (N=9,665), 42 (N=9,841) and 46 (N=8,851).

Add Health is a nationally representative cohort of ∼20,000 individuals in the US enrolled in grades 7-12 (aged 12-18) in 1994-1995 (19). The cohort has been followed up in four additional waves, with the most recent Wave V (N=12,297) occurring in 2016–18 (ages 33-43). Biomedical measures were collected on a subsample of participants at Wave V (N=5,381).

### Variables

Our outcome variables were smoking status; alcohol consumption; body mass index (BMI); self-rated health; cholesterol; blood pressure (BP, hypertension); and blood sugar level (glycated haemoglobin [HbA1c], a marker for diabetes). In BCS70, smoking status and alcohol consumption were measured at age 34, self-rated health at age 42, and all remaining measures at age 46. For Add Health, all measures were taken from Wave V. Outcomes were converted to binary variables using cut-offs shown in Supplementary Methods S1.

For chronic diseases (e.g., diabetes) measured through biomarkers (e.g., HbA1c), we distinguish between the biomarker alone, and “any” indication of the disease (e.g., reported as ‘any Diabetes’) which additionally draws on medication usage for the specific conditions.

For obesity, we distinguish between a measure based on measured height and weight, and a measure supplemented with self-reported height and weight (used in the main analysis). For alcohol, we define heavy drinking based on number of drinks, using US heavy drinking guidelines. Full details of the harmonisation of measures are shown in Methods S2.

Three measures of SEP were used: parental education (Add Health ages 11-19, BCS70 age 16), own education, and household income (Add Health ages 32-42, and BCS70 ages 34 and 42, dependent on the respective outcome). In both cohorts, parental education was grouped as: 1) neither parent has a university bachelor’s degree; 2) at least one parent has a degree. Own education was also grouped based on completing a bachelor’s degree. In both cohorts, own income was classified into approximate quintiles. For further details see Methods S2.

BCS70 is largely racially/ethnically homogenous, with most of the cohort born to parents who were themselves born in the UK or Europe (93%), and therefore likely White (see Methods S2 for further details). In Add Health, race/ethnicity was measured at Wave I; the primary analysis was restricted to non-Hispanic White adults to maximize comparability with BCS70.

In BCS70, the “age 46” biomedical sweep took place across 3 years (ages 46-48), while age in Add Health was calculated based on interview and birth date. In the remaining BCS70 sweeps, age was included as a dummy variable (i.e., 34 and 42). Sex assigned at birth was measured in the first sweep in BCS70 and at Wave I in Add Health.

### Statistical Analysis

Modified Poisson regression analysis was used to obtain relative risk estimates (risk ratios [RR]) and corresponding 95% confidence intervals (CI), using Stata 17. In Model 1, the independent variable was a dummy variable for country (Britain or US) controlled for age. Model 1 was run in both pooled and sex-stratified samples. In the sensitivity analysis, the full, racially/ethnically heterogenous Add Health sample was used.

Model 2 examined modification in the associations between early life SEP (parental education) and current adult SEP (education and household income) and health, including interaction terms between country and SEP. For interpretation, RR estimates are presented as adjusted predicted marginal estimates of prevalence for each country and/or for each country at each level of SEP, estimated at the observed values of covariates (further details are provided in Methods S3). A Wald test indicated whether SEP differences were significant between countries. For household income, the difference between the bottom, middle, and top quintile relative to the second quintile was tested for significance, controlled for household size.

Model 3 examined differences in the relationship between childhood SEP and adult health outcomes after controlling for adult education and household income. Models 2 and 3 were stratified by sex in supplementary analyses.

Sensitivity analyses compared models using clinically measured obesity in Add Health with the self-report supplemented measure used in the main analysis. Alternative harmonised measures of heavy drinking, based on UK and country specific guidelines, were also examined.

### Complex Survey Design and Non-Response Weights

Add Health uses a complex, stratified sampling strategy (19), thus maintaining the national representativeness of the data. Add Health also includes survey weights that account for non-representativeness among adults providing biomarker samples.

To ensure the complex survey design of Add Health and non-response was accounted for in analysis, non-response weights were developed in BCS70. The full method used to develop non-response weights, and to apply a complex survey design, is shown in Methods S4 & S5.

## Results

### Descriptives

Table 1 shows the weighted proportion of outcomes and covariates in the analytic samples (limited to non-Hispanic White in Add Health; unweighted distribution in Supplementary Results S1). In the US, there was higher prevalence of obesity, high blood pressure and cholesterol, whilst in Britain a higher proportion reported poor self-rated health, heavy drinking, and regular smoking. A higher proportion of the US sample had degree-level educated parents (36% versus 21%), whilst degree completion among cohort members was similar (40% versus 36%).

**Table 1.**
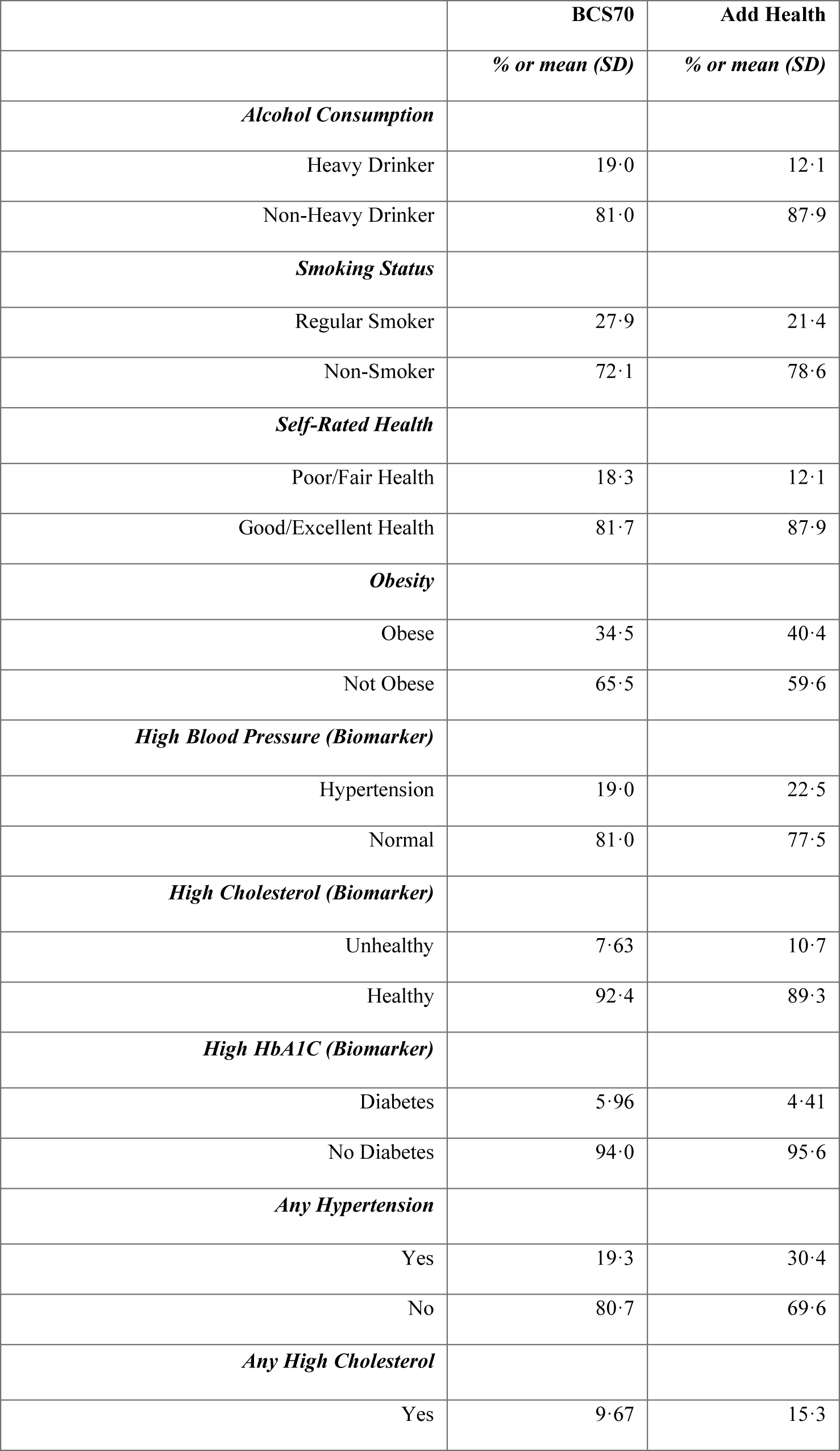
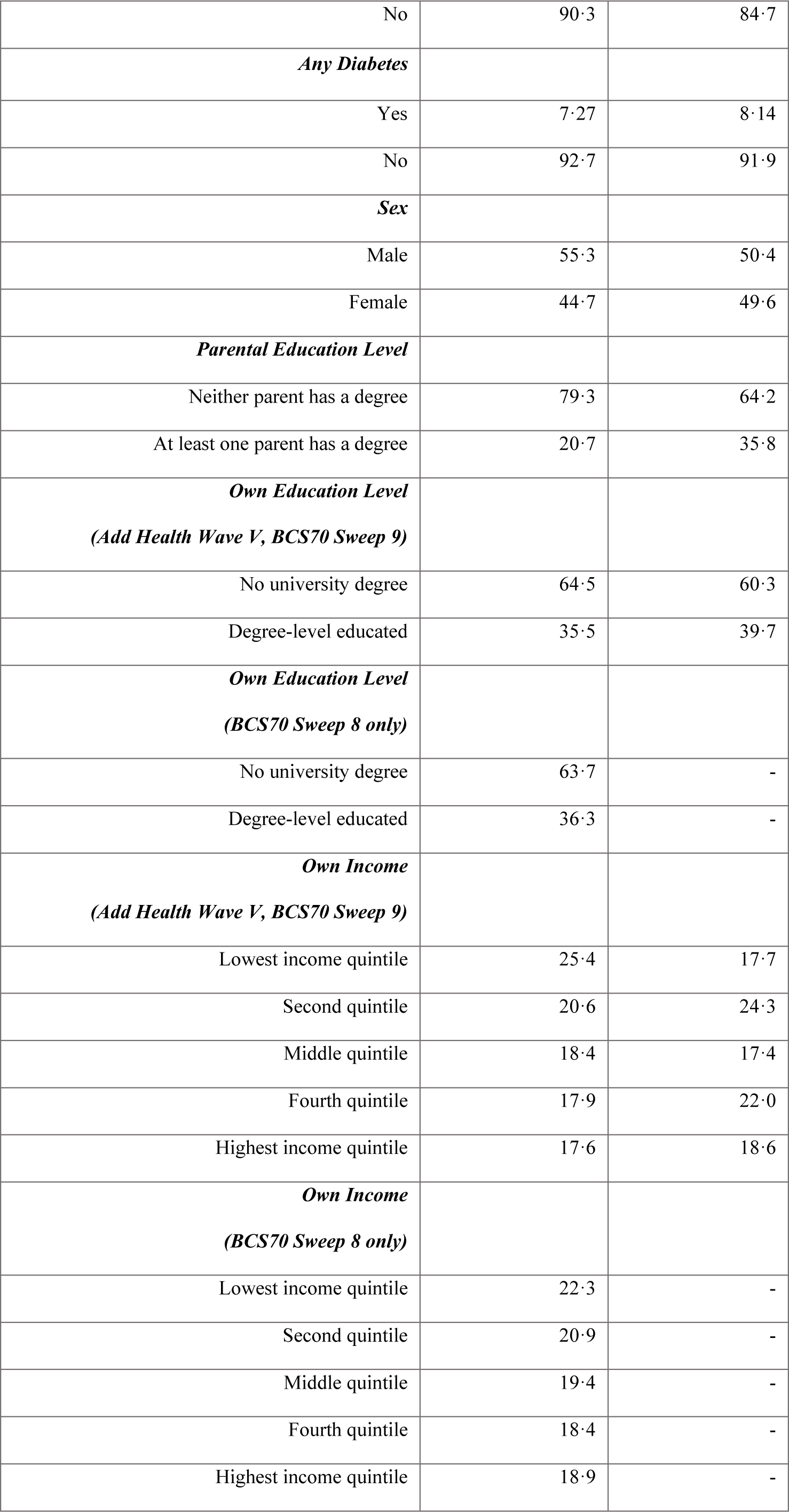
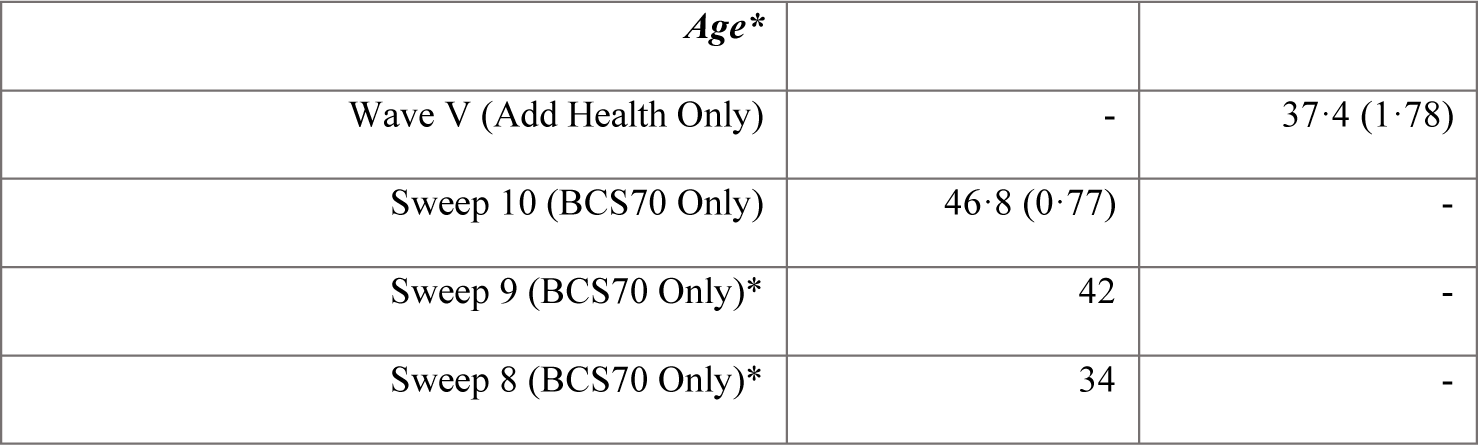
Weighted distribution of outcomes and covariates in BCS70 and Add Health in analytic sample (non-Hispanic White only). **Table 1 Footnote:** For all outcomes the values in the table represent weighted proportions (%), apart from age which represents mean and SD. * Age at sweep 9 and 8 in BCS70 is included as a dummy variable, for the respective age in years at interview. Therefore, standard deviation for these ages is equal to 0, as all cohort members are allocated the same age in years.

### Model 1 – Comparison of health between Britain and the US

US adults generally had worse cardiometabolic health than in Britain (Figure 1, Table 2). They were more likely to have high blood pressure and cholesterol, before and after accounting for medication use (any hypertension: 0·309 [95% CI: 0·284, 0·335] vs 0·193 [95% CI: 0·181, 0·204] ; any high cholesterol: 0·159 [95% CI: 0·138, 0·181] vs 0·097 [95% CI: 0·086, 0·107]), and more likely to have obesity (0·405 [95%CI: 0·384, 0·426] vs 0·345 [95%CI: 0·332, 0·358]). However, British adults were more likely to be smokers (0·279 [95%CI: 0·268, 0·290] vs 0·214 [95%CI: 0·195, 0·234]), heavy drinkers (0·190 [95% CI: 0·181,0·199] vs 0·121 [95% CI: 0·110, 0·131]) and to have poor self-rated health (0·183 [95% CI: 0·172, 0·194] vs 0·122 [95%CI: 0·108, 0·136]). They also had slightly higher HbA1c levels: 0·060 [95% CI: 0·051, 0·068] vs 0·044 [95% CI: 0·034, 0·055].

**Figure 1.**
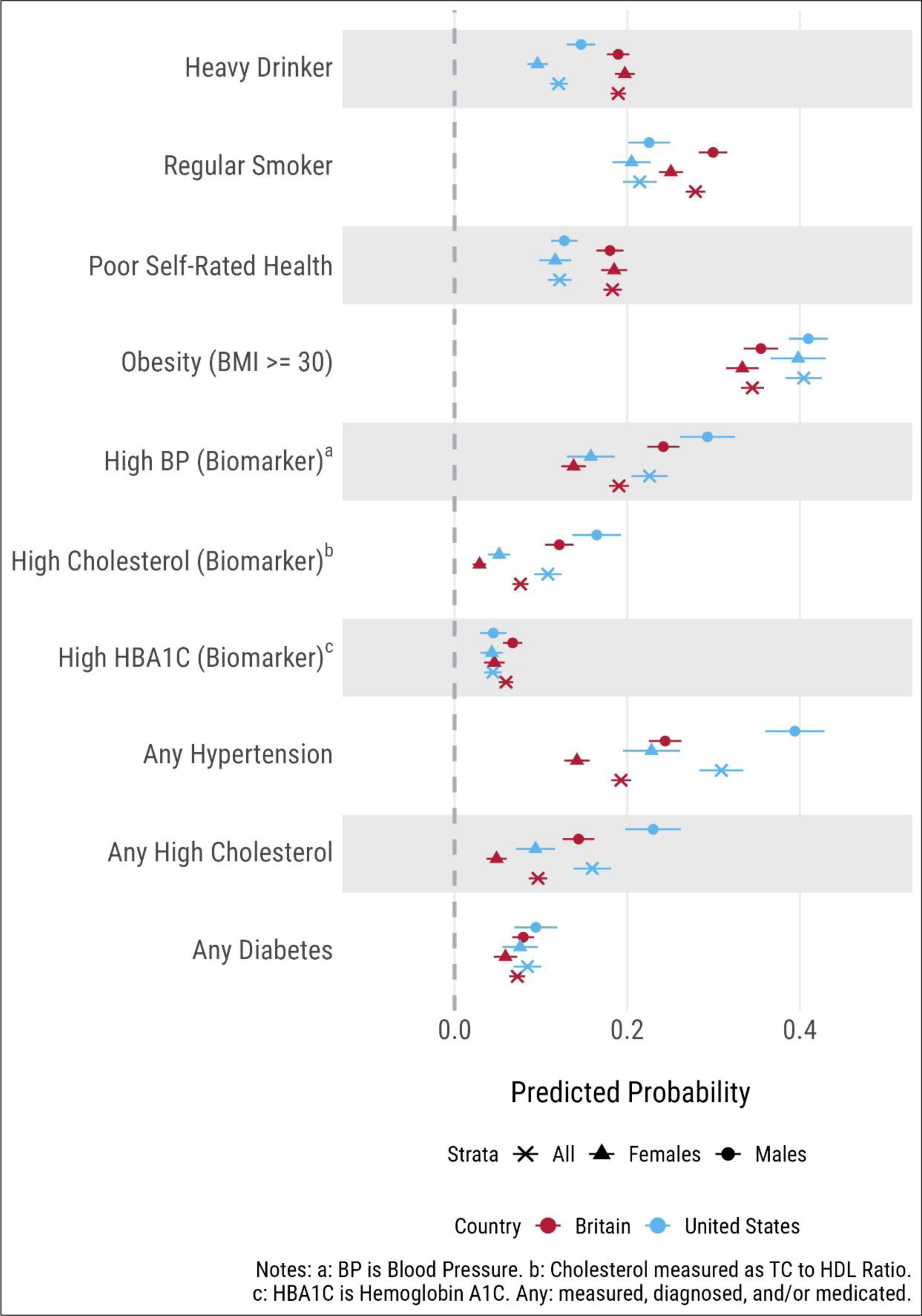
Predicted probabilities from modified Poisson regression, comparing health indicators between Britain and the US, by sex (Model 1). **Figure 1 Footnote:** Predicted probabilities from modified Poisson regression, comparing health indicators between Britain and the US in the whole sample and stratified by sex (Model 1). Outcomes labelled “any” refer to biomarkers that have been supplemented with medication use, therefore indicating a positive diagnosis of diseases.

**Table 2.** Marginal estimates from modified Poisson regression for Model 1, examining country differences in health outcomes: overall and sex stratified. **Table 2 Footnote.** Results presented are for model 1, exploring country differences in health outcomes between the UK and US. Note: BP is Blood pressure; high cholesterol measured by total cholesterol (TC) to High-density lipoprotein (HDL) ratio; HbA1c is Glycated haemoglobin (blood sugar levels). Outcomes labelled “any” refer to biomarkers that have been supplemented with medication use, therefore indicating a positive diagnosis of diseases. * P-value is for Wald test, indicating a statistical difference between countries.

|  |  | Britain |  |  | United States |  |  |  |
| --- | --- | --- | --- | --- | --- | --- | --- | --- |
|  |  | Estimates | Lower 95% CI | Upper 95% CI | Estimates | Lower 95% CI | Upper 95% CI | P Value<br>Difference* |
| *Heavy Drinking* | All | 0.190 | 0.181 | 0.199 | 0.121 | 0.110 | 0.131 | <0.0001 |
|  | Males | 0.190 | 0.177 | 0.203 | 0.147 | 0.130 | 0.163 | <0.0001 |
|  | Female | 0.197 | 0.186 | 0.209 | 0.096 | 0.084 | 0.108 | <0.0001 |
| *Smoking* | All | 0.279 | 0.268 | 0.290 | 0.214 | 0.195 | 0.234 | <0.0001 |
|  | Males | 0.299 | 0.283 | 0.316 | 0.225 | 0.201 | 0.250 | <0.0001 |
|  | Female | 0.251 | 0.237 | 0.264 | 0.205 | 0.183 | 0.227 | 0.0006 |
| *Self-rated Health* | All | 0.183 | 0.172 | 0.194 | 0.122 | 0.108 | 0.136 | <0.0001 |
|  | Males | 0.180 | 0.164 | 0.196 | 0.127 | 0.112 | 0.142 | <0.0001 |
|  | Female | 0.185 | 0.170 | 0.200 | 0.117 | 0.098 | 0.135 | <0.0001 |
| *Obesity* | All | 0.345 | 0.332 | 0.358 | 0.355 | 0.335 | 0.375 | <0.0001 |
|  | Males | 0.355 | 0.335 | 0.375 | 0.410 | 0.387 | 0.432 | 0.0003 |
|  | Female | 0.333 | 0.314 | 0.352 | 0.398 | 0.366 | 0.430 | 0.0006 |
| *High BP (Biomarker)* | All | 0.190 | 0.179 | 0.202 | 0.226 | 0.205 | 0.247 | 0.0035 |
|  | Males | 0.242 | 0.223 | 0.260 | 0.293 | 0.261 | 0.325 | 0.0066 |
|  | Female | 0.138 | 0.124 | 0.152 | 0.158 | 0.130 | 0.186 | 0.209 |
| *High Cholesterol*<br>*(Biomarker)* | All | 0.076 | 0.067 | 0.086 | 0.108 | 0.092 | 0.124 | 0.0006 |
|  | Males | 0.121 | 0.105 | 0.138 | 0.165 | 0.136 | 0.193 | 0.0094 |
|  | Female | 0.029 | 0.021 | 0.037 | 0.052 | 0.039 | 0.064 | 0.0028 |
| *High HbA1c*<br>*(Biomarker)* | All | 0.060 | 0.051 | 0.068 | 0.044 | 0.034 | 0.055 | 0.024 |
|  | Males | 0.067 | 0.056 | 0.079 | 0.045 | 0.029 | 0.060 | 0.0201 |
|  | Female | 0.046 | 0.034 | 0.058 | 0.043 | 0.030 | 0.056 | 0.75 |
| *Any Hypertension* | All | 0.193 | 0.181 | 0.204 | 0.309 | 0.284 | 0.335 | <0.0001 |
|  | Males | 0.244 | 0.225 | 0.263 | 0.394 | 0.360 | 0.429 | <0.0001 |
|  | Female | 0.142 | 0.127 | 0.156 | 0.228 | 0.195 | 0.261 | <0.0001 |
| *Any High Cholesterol* | All | 0.097 | 0.086 | 0.107 | 0.159 | 0.138 | 0.181 | <0.0001 |
|  | Males | 0.144 | 0.125 | 0.162 | 0.230 | 0.198 | 0.262 | <0.0001 |
|  | Female | 0.049 | 0.037 | 0.060 | 0.094 | 0.071 | 0.116 | 0.0005 |
| *Any Diabetes* | All | 0.073 | 0.063 | 0.082 | 0.084 | 0.068 | 0.100 | 0.21 |
|  | Males | 0.080 | 0.067 | 0.092 | 0.094 | 0.069 | 0.119 | 0.3006 |
|  | Female | 0.059 | 0.045 | 0.073 | 0.076 | 0.055 | 0.097 | 0.17 |

In Britain and the US, men were more likely to have high blood pressure and cholesterol compared to women. Men were also more likely to be heavy drinkers in the US and to be regular smokers in Britain. The male health disadvantage was greater in the US for heavy drinking and high cholesterol and blood pressure. In Britain, there was a larger male disadvantage for smoking.

### Model 2 – Socioeconomic inequalities between Britain and the US

Socioeconomic inequalities in midlife health were greater for adult SEP compared to childhood SEP in both cohorts (Figure 2, Results S2). The probability of being a regular smoker and reporting poor self-rated health was higher for individuals in the bottom income quintile, and for those without a degree. There were no differences in the probability of heavy drinking by SEP in the US, whilst more advantaged British adults (based on income and parents’ education) were more likely to be heavy drinkers.

**Figure 2.**
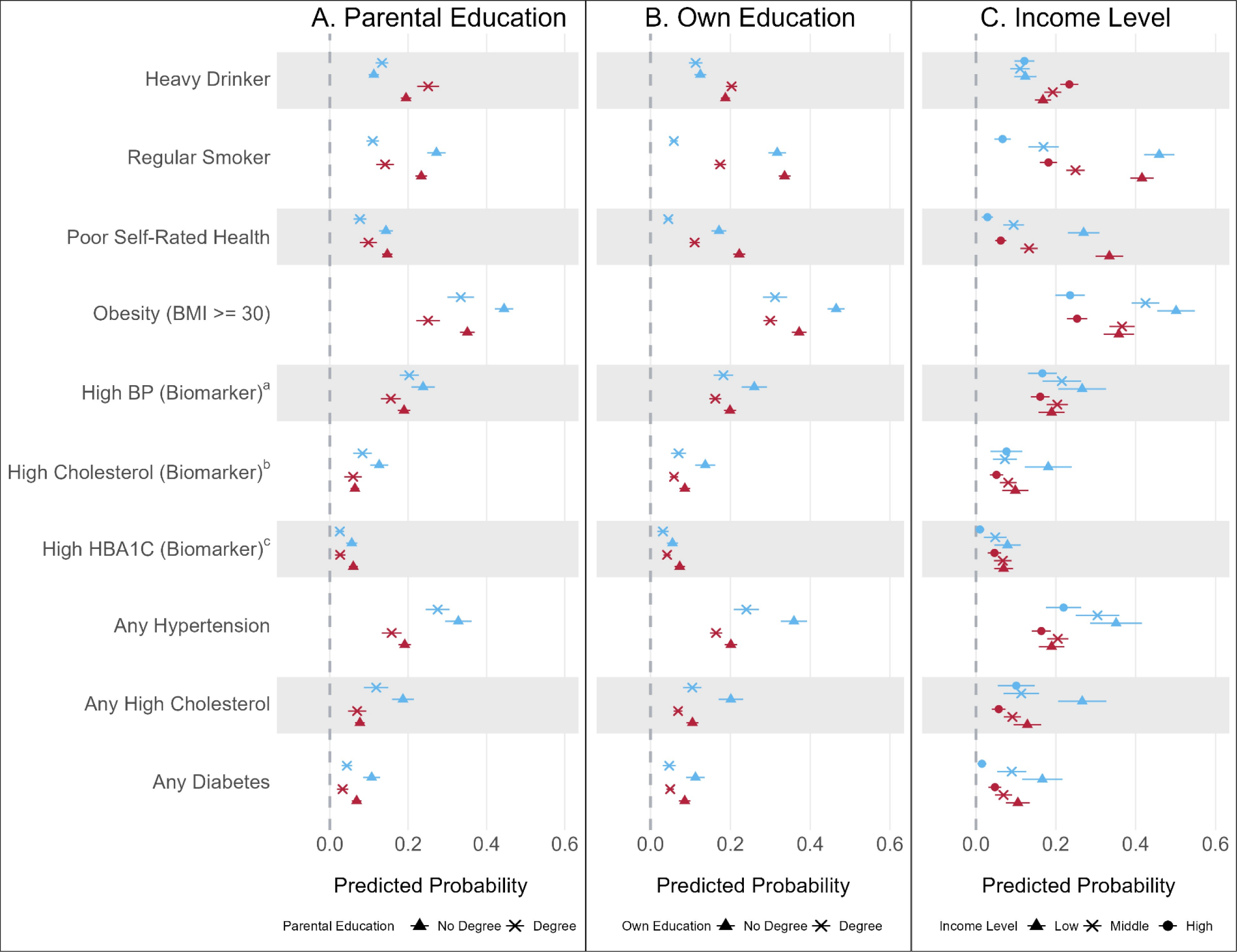
Predicted probabilities from modified Poisson regression showing socioeconomic inequalities in midlife health between Britain and the US (Model 2a, 2b and 2c). **Figure 2 Footnote:** Predicted probabilities from modified Poisson regression, comparing socioeconomic inequalities in health indicators between Britain and the US (Model 2). Measures of SEP are parental education (Model 2a), the cohorts own education level (Model 2b), and household income quintiles (Model 2c, only the first, third and fifth quintiles presented in the figure). Note: a) BP is Blood pressure; b) is cholesterol measured by total cholesterol (TC) to High-density lipoprotein (HDL) ratio; c) HbA1c is Glycated haemoglobin (blood sugar levels). Outcomes labelled “any” refer to biomarkers that have been supplemented with medication use, therefore indicating a positive diagnosis of diseases.

In Britain and the US there was a small SEP gradient in hypertension and cholesterol, but this was only significant for adulthood education. There was no difference between middle- and low-income groups in the probability of obesity in Britain (Figure 2 Panel C, 0·366 [95%CI: 0·334, 0·397] vs 0·358 [95%CI: 0·320, 0·396]), but there was a difference between the highest and middle/lowest income quintiles. In the US there was a clearer income gradient for obesity (Panel C, low: 0·501 [95%CI: 0·454, 0·549], middle: 0·425 [95%CI: 0·390, 0·459], top: 0·236 [95%CI: 0·199, 0·273]). Both countries had gradients by education, which were stronger for participants’ rather than parent’s education. Results were similar when looking at males and females separately (Results S3).

For some outcomes, such as smoking, the most socioeconomically advantaged US adults were healthier than their British peers, but the most disadvantaged were worse off than the equivalent group in Britain, resulting in wider inequalities (Figure 2, Panel C, smoking. US top income: 0·067 [95%CI: 0·046, 0·087]; Britain top income: 0·182 [95%CI: 0·160, 0·203]; US bottom income: 0·459 [95%CI: 0·421, 0·497]; Britain bottom income: 0·416 [95%CI: 0·387, 0·445]). This was observed to a lesser extent for income differences in “any” diabetes.

For other outcomes, such as obesity, levels were similar among advantaged groups in Britain and the US, whilst disadvantaged US adults had higher obesity levels than in Britain, resulting in wider inequalities. Conversely, wider inequalities in self-rated health were the result of better self-rated health among advantaged US adults relative to Britain, despite similarly poor health among disadvantaged groups.

Finally, for some outcomes, such as hypertension and cholesterol, those in the most advantaged position in the US had similar or worse health than disadvantaged groups in Britain, particularly as measured by parental or adult education (Figure 2, Panel C, own education level and “any” hypertension. US has a degree: 0·240 [95%CI: 0·209, 0·272]; Britain does not have a degree: 0·164 [95%CI: 0·149, 0·179]). This pattern was also observed for obesity and parental SEP.

### Model 3 – Attenuation of associations with early life SEP

Model 3 examined associations between childhood SEP and adult health, controlling for adult SEP (Figure 3). Compared with Figure 2 Panel A, the associations between childhood SEP and most health outcomes were attenuated. However, this was not the case for smoking, where differences remained in the US. Conversely, inequalities remained largely unchanged in Britain for heavy drinking when accounting for adult SEP, compared to no inequality in the US. Results were similar when looking at males and females separately (Results S4), except for inequalities in HbA1c among males in Britain.

**Figure 3.**
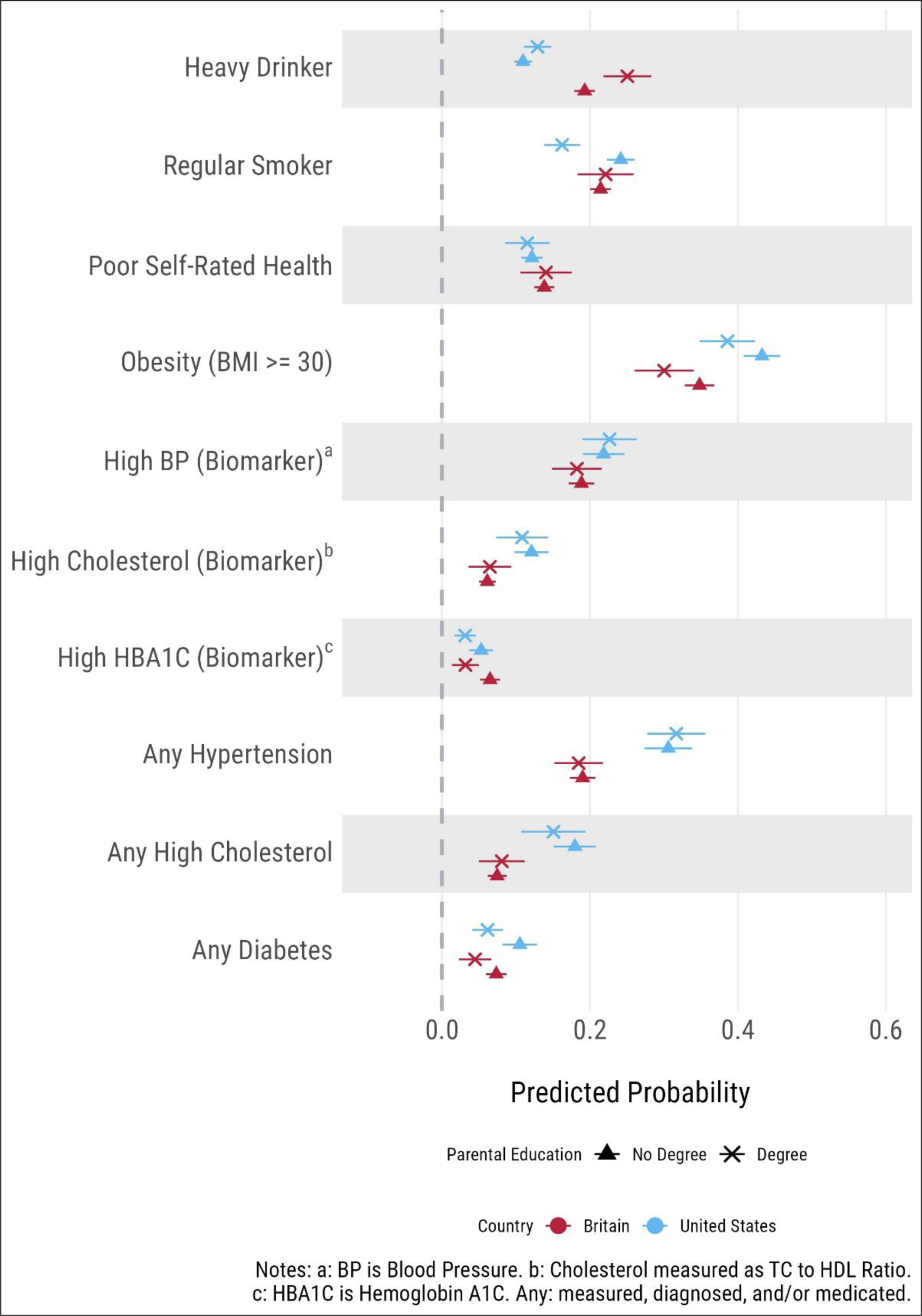
Predicted probabilities from modified Poisson regression of associations with parental education, adjusted for cohort members own SEP (education level and household income) in adulthood (Model 3). **Figure 3 Footnote:** Predicted probabilities from modified Poisson regression of associations with parental education, making adjustment for cohort members own SEP (education level and household income) in adulthood (Model 3). Outcomes labelled “any” refer to biomarkers that have been supplemented with medication use, therefore indicating a positive diagnosis of diseases.

### Sensitivity Analysis

Results using the full racially and ethnically diverse Add Health sample were similar to the main analysis (Results S5), with the exception of “any” diabetes, which was significantly higher in the US. In general, the gap between the two countries was smaller for heavy drinking, smoking, and self-rated health, but larger for obesity, high blood pressure, cholesterol, and blood glucose.

The results were also consistent when limiting obesity to directly measured height and weight in Add Health (Results S6). While the overall prevalence of obesity was higher in the US based on clinically assessed measures, conclusions were unchanged.

Finally, different operationalisations of heavy drinking (Methods S2 and Results S7) show that a US definition of heavy drinking based on number of alcoholic drinks results in a higher prevalence of heavy drinking in Britain, with SEP inequalities observed in Britain only. Comparatively, using a UK definition based on units, levels of heavy drinking were similar between Britain and the US with little evidence of inequalities in drinking behavior (except own education in the US). When using country specific definitions (based on UK guidelines for units in BSC70, and US guidelines for number of drinks in Add Health) – therefore capturing excess drinking based on country specific guidelines– heavy drinking was more common in the US compared to Britain, with little evidence of inequalities in either cohort.

## Discussion

Our analyses identified a US health disadvantage at midlife similar to that observed at older ages (1–4). The health disadvantage is notable for obesity, high blood pressure and cholesterol, whilst Britain exhibits greater prevalence of smoking, heavy drinking, and worse self-rated health. Our results demonstrate that socioeconomic inequalities are typically wider in the US, where health differences between the most and least advantaged are larger. For smoking, and to a lesser extent diabetes, this is due to better health among the most advantaged in the US but worse health among the most disadvantaged compared to Britain. For hypertension and high cholesterol, the most advantaged adults in the US have health that is equivalent to (or worse than) the most disadvantaged adults in Britain. In both countries, socioeconomic inequalities in health were typically wider for adult SEP than parent’s SEP; likewise, most of the associations with parental SEP were attenuated by adult SEP.

Our finding that hypertension and high cholesterol are more frequent in the US supports previous research documenting worse cardiometabolic health among older US adults (2, 4). While research on midlife is more limited, our results also find a US disadvantage with respect to obesity and cardiometabolic health (5) but differ from work suggesting worse hypertension and dyslipidaemia among more recent midlife cohorts in England (10). Sensitivity analyses found that the risk of diabetes in the US was higher only when considering the full ethnically heterogenous sample in Add Health. This finding differs from previous work documenting double the diabetes prevalence among US older adults compared to England (2), even in a non-Hispanic White sample, suggesting a possible increase in diabetes risk among younger British cohorts (10). Nevertheless, we find substantial evidence of poor health in midlife among both cohorts, supporting prior literature on declines in midlife health, such as higher prevalence of obesity, psychological distress and multimorbidity in Britain (20–22), and increases in midlife mortality in the US (23). Indeed, our work highlights the importance of studying healthy ageing as a process that occurs across the whole lifespan (7, 24).

Previous work found the co-occurrence of risky behaviours, including smoking and drinking (1, 5), to be more common among older and middle-aged adults in England than the US. This is consistent with the higher prevalence of unhealthy behaviours in Britain compared to a higher burden of chronic disease risk in the US observed in our study. This seeming contradiction suggests that the US health disadvantage is attributable to a multitude of mechanisms (e.g., diet, physical activity, and other health-relevant lifestyle factors) that future research should examine.

However, we caution that the harmonization of self-reported measures of health remains a challenge in internationally comparative research, where the subjective nature of both interpretation and reporting is important to account for. For example, in supplementary materials (Results S7, Discussion S1) we show how results differ based on the operationalization of heavy drinking, and the extent to which more or less conservative definitions and cutoffs might capture different ends of the distribution of drinking behaviours, in turn driving observed differences between countries and SEP gradients.

For several outcomes we find that the most socioeconomically advantaged respondents in the US have equal or worse health compared to the most disadvantaged in Britain. This may reflect different sociopolitical contexts between the two countries. For example, the US and UK health care systems differ substantially (25). The UK has the National Health Service, which is universally available and free at the point of access. In the US, healthcare is largely covered by private health insurance, Medicare or through an individual’s own finances, and the associated costs are often high. Past work has suggested that relatively “universal” access to healthcare at older ages in the US through Medicare helps explain its better international standing in mortality and morbidity for medically amenable causes of death (26). However, comparisons of income gradients in health at older ages (70-80 years of age) between England and the US found no differences in England compared to a clear gradient in the US. As the authors note, this is likely due to a more generous benefit system for older adults in England where, below the median income, retirement benefits are largely consistent and unrelated to historic income (3).

It is also possible that differences between the US and Britain reflect broader inequalities affecting health across the life course. Societies with higher levels of inequality perform worse in international comparisons across a range of health indicators (27). The impact of socioeconomic inequalities within each country may be relevant to the present study’s finding of worse health in the US, where the unique combination of high inequality and a weak welfare state in the US proves particularly harmful across *all* groups.

### Strengths and Limitations

This research utilises data from two nationally representative cohort studies in Britain and the US, exploring health differences on a range of outcomes, including physical measurements. BCS70 is representative of the population at time of recruitment; however, most of the cohort are White and it was thus not possible to make similar adjustments for race/ethnicity in both studies. Also, for biomedical measures, the age at which measures were collected did not fully overlap between Add Health and BCS70. Moreover, there likely remains residual confounding for associations with SEP across the outcomes considered.

Our extensive harmonisation of measures between the two cohorts included the development of novel weights in BCS70, allowing for comparative analysis that accounts for Add Health’s complex survey design. Despite efforts to address attrition through use of non-response weights, the derivation of weights was not identical, and some residual bias remains.

As is the case with harmonisation, there may be residual differences in how variables were measured and understood. This is especially problematic for subjective measures such as smoking, drinking, and self-rated health, where questions are asked and/or possibly interpreted differently. This work highlights a need for better international harmonisation of longitudinal studies at younger ages given the success of previous efforts in providing evidence on health disparities at older ages.

## Conclusion

We find that US adults in midlife have worse cardiometabolic health than those in Britain, as well as wider SEP inequalities for multiple health outcomes. For some cardiometabolic outcomes, even the most advantaged SEP groups in the US have worse health than all groups in Britain. Our work also highlights the need for more efforts to harmonise international datasets at younger ages, and additional research that explores specific contextual drivers of international differences in health and inequalities in midlife.

## Data Availability

All data in BCS70 is available through an end user license through UKDS: https://beta.ukdataservice.ac.uk/datacatalogue/series/series?id=200001. Add Heath data can be accessed through a data application, see further details here: https://addhealth.cpc.unc.edu/data/.

https://beta.ukdataservice.ac.uk/datacatalogue/series/series?id=200001

## Author Contribution

CBS and IG contributed equally to manuscript, and therefore have the right to list themselves as first author when presenting or using this work. CBS was responsible for data preparation, harmonisation of variables across the cohorts, derivation of non-response weights in BCS70, writing, preparation and reviewing of the manuscript. IG was responsible for data preparation, harmonisation of variables across the cohorts, running the analysis, writing, preparation and reviewing of the manuscript. AT was responsible for creation of figures, writing, preparation and reviewing of the manuscript. LGi, MN and VM contributed to development of code and/or harmonisation of variables and reviewing of the manuscript. GP, JBD and LGa are the senior authors on this paper, who were responsible for project development, supervision and reviewing of the manuscript. All authors contributed to initial conceptualisation and design of the research and reviewed the manuscript.

## Data Sharing

All data in BCS70 is available through an end user license through UKDS: https://beta.ukdataservice.ac.uk/datacatalogue/series/series?id=200001. Add Heath data can be accessed through a data application, see further details here: https://addhealth.cpc.unc.edu/data/. All code associated with the current analysis can be found in the following OSF repository: https://osf.io/vkf2g/?view_only=89368400bf79432581a66fc681d914e7

## Acknowledgements

We thank the study participants for their continued involvement in BCS70 and Add Health, as well as CLS colleagues who contributed to early discussions around this work (Jenny Chanfreau and Alice Goisis). We are grateful for feedback provided on previous versions of this work, that were presented at the British Society for Population Studies conference in September 2023, the Interdisciplinary Association for Population Health Science in October 2023, and the Society for Longitudinal and Lifecourse Studies in October 2023.

