## Supplementary Material for "Midlife Health in Britain and the US: A comparison of Two Nationally Representative Cohorts"

### **Supplementary Methods S1: Outcome cut-offs used in BCS70 and Add Health**

**Supplementary Methods Table 1.** Description of health behaviour, anthropometry and biomarker outcomes and the cut-offs used in BCS70 and Add Health

| **Outcomes** | **Cut-off used** |
| --- | --- |
| Smoking | 1) “Regular smoker (smokes a cigarette every day)” versus 2) “Occasional smoker, past smoker or never smoked (Does not currently regularly smoke cigarettes)” |
| Alcohol consumption ^A^ | Heavy drinker (Men): > 15 drinks a week Heavy drinker (Women): > 8 drinks a week |
| Self-rated health | 1) “Poor, or fair health” versus 2) “Good, very good, or excellent health” |
| BMI ^B^: Obese | Obesity: >=30 kg/m^2^ |
| Cholesterol: Total/HDL ratio | Unhealthy Levels: >=6 |
| Glycated haemoglobin: HbA1c | Diabetes: 6·1% (>= 43·2 mmol/Mol) |
| Blood Pressure | Hypertension: Systolic >=140 or Diastolic >=90 ^C^ |

***Supplementary Methods Table 1 Footnote:*** *^A^ Heavy drinking was measured as units per week in BCS70, and number of alcoholic drinks per week in Add Health. ^B^ Body Mass Index (BMI) was derived from objectively measured height and weight in both cohorts but supplemented with self-reported height and weight where nurse/examiner measured was unavailable. ^C^ Respondents were classified as having hypertension if either systolic or diastolic blood pressure was above the cut-off.*

### **Supplementary Methods S2: Harmonisation of measures between Add Health and BCS70**

Smoking

In BCS70, respondents were asked at age 34 to identify on a card whether they would say that they had “never smoked cigarettes”, that they “used to smoke cigarettes but don’t at all now”, that they “smoke cigarettes occasionally but not every day”, or that they “smoke cigarettes every day”.

In Add Health, respondents were asked in wave 5 how many days out of the last thirty days they had smoked cigarettes, with possible responses ranging from 0 to 30 days.

Using these answers to these questions, we identified current regular cigarette smokers and those who did not regularly smoke cigarettes (see below, Supplementary Methods Table 2).

***Supplementary Methods Table 2i.*** *Categorisation of smokers by responses in Add Health and BCS70.*

|  | **BCS70** | **Add Health** |
| --- | --- | --- |
| **Current regular smoker.** | “Smoke cigarettes every day” | In the last 30 days, smoked cigarettes on 30 days |
| **Does not currently regularly smoke cigarettes.** | “Smoke cigarettes occasionally but not every day” **OR** “Never smoked cigarettes” **OR** “Used to smoke cigarettes but don’t at all now” | In the last 30 days, smoked cigarettes on <30 days |

Alcohol consumption

In BCS70, cohort members self-reported their drinking habits in the last seven days at age 34 by reporting the number of drinks they had across a range of different alcoholic drink types (e.g., beer, wine, spirits, alcopops, fortified wine). For beer, the question worded in terms of number of units consumed (i.e. “Number of units of beer within the last seven days”), therefore equivalent to one half pint. For other types of drinks, reports for different numbers of drinks consumed were combined and converted into alcohol units per week (based NHS guidelines for alcohol units in different drinks: https://www.nhs.uk/live-well/alcohol-advice/calculating-alcohol-units/).

For Add Health, the variables used were how many days per month the cohort member drinks, as well as how many drinks the cohort member has each time they drink, which were recoded and combined to reflect the number of drinks per week. In Add Health, no distinction is made between types of alcoholic drinks (e.g., beer or wine).

Different cut-offs for heavy drinking exist in Britain and the US, with drinking guidelines based on units in Britain and number of drinks in the US. In Britain, heavy drinking is defined using the UK National Institute for Health and Care Excellence cut-offs (National Institute for Health and Clinical Excellence. Alcohol-use disorders: diagnosis and clinical management of alcohol-related physical complications. (Clinical guideline CG100.) 2010. http://guidance.nice.org.uk/CG100), with heavy drinking among men defined by consuming more than 50 alcohol units a week, and more than 35 alcohol units a week among women. In the US heavy drinking is defined according to the Centers for Disease Control and Prevention (CDC) with heavy drinking among men defined as consuming 15 drinks or more a week, and 8 drinks or more a week among women.

Therefore, four different approaches were explored in sensitivity analysis to harmonise alcohol consumption between the cohorts (conversion between drinks and units was done based on one-unit equalling 10ml or 8g of pure alcohol, and where a standard drink in the US contains 14g of pure alcohol):

1) Adopting the country specific definitions for heavy drinking within the respective cohorts (e.g., using the unit-based cut-off in BCS70 and the drinks-based cut-off in Add Health). This variable would therefore assess relative excess drinking based on culturally specific norms, rather than an absolute comparison of consumption of alcoholic drinks or units.
2) Converting units in BCS70 to number of drinks (multiply by 0·57) and adopting the US drink-based cut-off for heavy drinking in both cohorts.
3) Use the number of drinks as originally reported in BCS70 (only converting beer into number of drinks) and adopting the US drink-based cut-off for heavy drinking in both cohorts.
4) Converting number of drinks in Add Health to units (multiply by 1·75) and adopting the British unit-based cut-off for heavy drinking in both cohorts.

For the main analysis, we use version 3 of the harmonised variables for alcohol consumption, therefore using number of drinks as originally reported in BCS70 and adopting the US definition of heavy drinking. This measure was chosen of the four to be included in the main analysis, as it requires the fewest assumptions to be made when creating a harmonised measure that attempts to compare absolute consumption of alcohol. However, there are still limitations to this approach, as no description of the type of drink is provided in Add Health, so assumptions are still made about the comparability of “number of drinks” between the cohorts.

Self-rated health

In BCS70 at age 42, participants were asked to describe if their health was 1) Excellent; 2) Very good; 3) Good; 4) Fair; and 5) Poor. Cohort members were then grouped into those who described their health as "Excellent/Very Good/Good" and "Fair/Poor".

Similarly, in Add Health in Wave V, participants were also asked to describe their health on this five-point scale, and their responses were grouped using the same categorization schema as above.

BMI: Obesity

In BCS70 at age 46, weight in kilograms (kg) was collected through Tanita BF-522W scales. The scales can accurately measure up to 130kg, and only those whose weight was unlikely to exceed this amount were weighed. Height was measured by nurses using a portable Leicester stadiometer following standard protocol. Cohort members also self-reported height and weight.

Body mass index (BMI) was derived by dividing nurse measured weight in KG by nurse measured height in meters (m) squared (kg/m^2^). Where nurse measured BMI could not be derived, BMI using self-reported height and weight was derived instead using the same method (n= 1,081), and supplemented the nurse measured BMI (n= 7,413). Individuals with a BMI exceeding or equal to 30 kg/m^2^ were categorised as obese, whilst individuals with a BMI lower than this threshold were categorised as not obese.

Likewise, in Add Health at Wave V weight and height were also collected at time of examination among the subset of respondents participating in the biomarker specimen collection (n=5,377). Add Health provides users with both calculated BMI and categorized BMI for these respondents, using the same 30 kg/m^2^ threshold for obesity. Supplementary information on individuals’ BMI was obtained from self-reported height and weight (n=6,839), which is asked of all respondents in the survey component of the data collection.

Cholesterol: Total/HDL ratio

In BCS70, cohort members provided a blood sample at age 46 and total cholesterol and HDL cholesterol were measured. Cohort members were categorised as having high cholesterol (hyperlipidaemia) if the ratio of total cholesterol to HDL cholesterol exceeded 6.

Additionally at age 46, nurses looked at medical records of a subset of the sample and reported medications being used by the cohort members, including lipid regulating medication. If cohort member were using lipid regulating medication but there total/HDL ratio was “healthy”, they were reclassified as having high cholesterol.

In Add Health, cohort members provided a blood sample at ages 33-43 (varying across members) and total cholesterol and HDL cholesterol were measured. The same cut-off (total cholesterol to HDL cholesterol ratio greater than 6) was used to categorise hyperlipidaemia.

Additionally, Add Health respondents provided self-reported information on their medication use, including lipid regulating medication. This information was collected in both the in-home survey (respondents who first reported receiving a diagnosis for hyperlipidaemia were then asked if they take medication for their condition) and as part of the in-home examination for a subset of the sample (respondents were asked about what medications they take, which were then classified by Add Health staff). If respondents reported taking medication in response to either question, they were classified as having high cholesterol.

Glycated haemoglobin: HbA1c

In BCS70, glycated haemoglobin was measured when cohort members provided a blood sample at age 46 and were classified into those with diabetes if HbA1c levels exceeded 6·1% ( >= 43·2 mmol/Mol). Similar to the approach for cholesterol, those using diabetic medication as identified by nurses, were reclassified as having diabetes even if HbA1c levels were below the diabetic threshold.

In Add Health, cohort members provided a blood sample at ages 33-43 (varying across members) and were classified into those with diabetes based on the same HbA1c cut-off as in BCS70. As with high cholesterol, diabetic medication use was assessed in the in-home survey and examination, wherein either response led to respondents being classified as having diabetes.

Blood Pressure

At age 46 in BCS70 three blood pressure readings for systolic and diastolic blood pressure were collected. The mean was taken across the three readings, and a single blood pressure variable was derived if either the systolic or diastolic value exceeded their given threshold (Systolic >=140, Diastolic >=90). Individuals who were also reported by nurses to be taking medication for “hypertension or heart failure” at age 46 were reclassified to have high blood pressure (hypertension). Similar to other biomarkers, cohort members were also reclassified based on medication use if they did not have a blood pressure reading.

In Add Health, cohort members received a blood pressure reading for systolic and diastolic blood pressure. As with BCS70, the mean was taken across the three readings and a single blood pressure variable was derived if either the systolic or diastolic value exceed the aforementioned threshold. Once again, self-reported medication use in the in-home survey and examination portions of the survey was used to assess hypertension not captured by the blood pressure reading. Specifically, respondents were asked about hypertension and heart failure medication, separately, in the in-home survey; this was complemented by an Add Health derived indicator of antihypertensive medication use based on the examination questionnaire. Medication use related to hypertension and/or heart failure across these two sets of measures led to respondents being reclassified as having high blood pressure.

Childhood SEP: Parental Education

In BCS70, parental education was collected at age 16 through the family-follow up form, administered either by a health visitor during a home visit, or by self-completion sent in the post to be completed by the parent(s). The form ascertained if the mother, father, both parents or neither parent had the following qualifications: 1) trade apprenticeship or other occupational training; 2) O levels or equivalent (CSE/C&G etc); 3) A level or equivalent (OND/ONC/C&G); 4) nurse (SEN or SRN); 5) teacher (certificate of education or equivalent); 6) degree, diploma, or member of professional institute; 7) other qualification(s); 8) no qualifications; 9) qualifications of parents not known. From this, the highest education level of the mother and father were derived separately, and then combined to indicate if: 1) at least one parent has a degree or equivalent; or, 2) neither parent has a degree.

In Add Health, parental education was collected at Wave 1 when cohort members were aged 11-19. Both the cohort member and the parents were asked for the mother and fathers’ highest level of completed education. Responses were regrouped into those who 1) had a degree or equivalent (either “Graduated from college/university” or “Professional training beyond 4-year college”), and 2) no degree or equivalent (all other responses). Similar to BCS70, qualifications were combined between parents using the parent reported qualifications, to indicate if either parent had a university degree or if neither parent did. This was then supplemented with the cohort members response in the cohort member questionnaire, where parents own report of highest qualification was missing.

Adulthood SEP: Own Education

In BCS70, own education was assessed at age 34 and 42 by asking cohort members to report all academic and vocational qualifications obtained since their last interview. These were grouped into the National Vocational Qualification (NVQ) groupings of NVQ level 1-5 as derived by the BCS70 survey team, and a variable for Highest NVQ level from an academic or vocational qualification at the respective sweep was derived using prior survey data (for further details, please see: <https://cls.ucl.ac.uk/wp-content/uploads/2018/06/Deriving-highest-qualification-in-NCDS-and-BCS70.pdf> ).

For the present analysis, using the deposited derived variable for Highest NVQ level from an academic or vocational qualification, a new binary variable was derived grouping highest education level into 1) has a degree (e.g., NVQ level 4 & 5) and 2) does not have a degree (No qualification and NVQ level 1-3).

In Add Health, education of the cohort member was taken at Wave 5 when cohort members were age 33-43. Cohort members were asked to report their highest educational qualification achieved to date, and this was used to group cohort members into those who 1) have a degree (any response from the following: “completed college (bachelor's degree)", "some graduate school", "completed a master's degree", "some graduate training beyond a master's degree", "completed a doctoral degree", "some post baccalaureate professional education (such as law school, medical school, nursing)" and "completed a post baccalaureate professional degree (such as law, medicine, nursing)") and 2) those who do not have a degree (all other responses).

Adulthood SEP: Household Income

In BCS70 household income was assessed at ages 34 and 42 through the employment and family income questionnaire, which ascertained multiple sources of income as well as employment status for both the main respondent and partner. Household income was derived from the main respondent’s income, the partners income and any income from benefits or other sources whilst taking into account employment status. Full code and methodology for derivation of the income variable will be deposited with the UK data service at a later date. This was then classified into quintiles, resulting in five groups from the lowest income quintile to the highest income quintile.

In Add Health household income was taken from Wave 5 when cohort members were age 33-43. Cohort members were asked “What was the total household income before taxes and deductions in the last calendar year for all household members who contribute to household expenses?” and responded by indicating which income band they fell in, out of 13 bands ranging from “less than $5,000” to “$200,000 or more”. To allow comparability with BCS70, these were then regrouped into approximate quintiles, with roughly 20% of the respondents falling into each group. However, because these quintiles were being regrouped from banded categories, the proportion in each quintile ranged from 17·16% to 24·74%.

Ethnicity

BCS70 is nationally representative at the time of birth; due to few non-white births in Britain at the time of recruitment, BCS70 is predominantly an ethnically homogenous cohort.

In the BCS70 birth sweep, 95% of mothers and 94% of fathers of cohort members were themselves born in the UK or Europe. However, direct questions on the parents or cohort members ethnicity were not asked at the birth sweep. Therefore, it is not possible to conclusively identify cohort members’ ethnicity, as the ethnicity of the parent may not be the same as the majority ethnic group of the country they were born. At age 5, the interviewer reported the ethnicity of the parent being interviewed (mostly the mother), with 96% of respondents considered of UK or other European ethnicity. Among cohort members with data available on the biomarkers that are considered in the current analysis at age 46 (cholesterol, hypertension, and glycated haemoglobin), between 3·2% and 3·9% of the sample had a parent born outside of the UK or Europe. Therefore, ethnicity is not considered in analysis of BCS70.

In Add Health, ethnicity was measured at Wave I, and grouped as non-Hispanic White, Hispanic or Spanish/Latino, Black or African American, American Indian or Native American and Asian or Pacific Islander. For the current analysis, we distinguish between non-White and non-Hispanic White participants to aid comparison with BCS70.

***Supplementary Methods Table 2ii.*** *Unweighted and weighted distribution of Add Health sample by Ethnicity.*

| ***Ethnicity***  ***(Add Health only)*** | **N** | **Unweighted Proportion*** | **Weighted Proportion*** |
| --- | --- | --- | --- |
| Non-Hispanic White | 6,727 | 56·0 | 65·0 |
| Non-White and Hispanic White | 5,289 | 44·0 | 35·0 |
| Total | 12,016 | 100·0 | 100·0 |

***Supplementary Table 2ii Footnote: *****There is a discrepancy in the unweighted and weighted proportion of non-White and Hispanic Whites due to oversampling of minority ethnic groups in Add Health. However, this discrepancy is accounted for when weights are used in our main analysis.*

Age

In BCS70, age was recorded in years, up to the date of interview at the biomedical sweep at age 46, as data collection took place across 3 years, and therefore cohort members were aged between 45 and 48 at time of data collection. For age recorded at sweep 7 and sweep 9, age was included as a dummy variable, as 34 years of age and 42 years of age respectively. In Add Health, age was recorded in years at the time of data collection.

### **Supplementary Methods S3: Justification for analytic approach**

To help with the interpretation and visualisation of results from the non-linear models (1) and to allow for formal comparisons of differences (2), RR estimates were presented as adjusted predicted marginal estimates of prevalence for each country and/or for each country at each level of SEP, estimated at the observed values of covariates (i.e., the coefficients and error variances differ by country, rather than using representative or mean values). As is well established in extant literature on interaction terms in non-linear models (3), the significance of the interaction term does not provide conclusive evidence on country and/or SEP differences, as in this analysis. Rather, a formal comparison of marginal estimates and their corresponding 95% confidence intervals – which we examine using a Wald test of either the difference between countries (i.e. Model 1) or the SEP difference between countries (i.e. Models 2 and 3) – takes into account the country-specific observed values of the variables at baseline and the additional effect of the interaction term. Thus, while we include an interaction in our models, following best practices for non-linear models, we focus on marginal estimates where both additive and multiplicate effects are accounted for.

Further, this approach was used to allow for accurate weighted analysis, using non-response weights in Add Health that allow for national representativeness, and those developed in BCS70. The scales of the weights in the two samples are very different. Specifically, Add Health respondents are weighted such that each individual represents the corresponding number of adults in the population that share the sociodemographic criteria used as part of the sampling (while accounting for nonresponse); thus, the sum of these weights approximates the size of the U.S. cohort from which Add Health respondents were sampled. The BCS70 weights do not use this approach, as the sample is broadly representative of the British population born in 1970 without this adjustment. Due to the unique, survey-specific derivation of these weights, the distribution of covariates in each dataset needs to be accounted for in the analysis as to not yield inaccurate estimates for *both* samples. Additionally, as our analysis looks at two different populations, means in covariates across populations do not exist.

Inequalities were therefore evaluated on the country specific SEP average. This allows inequalities to be assessed both in absolute and relative terms. E.g.:

- Relative inequalities perspective: people are subject to SEP inequalities in health based on the magnitude of inequality *where they live*, regardless of the baseline.
- Absolute inequalities perspective: income may mean different things with respect to purchasing power or status, among other examples, as is true for returns to education, such that equivalent levels of income and/or education are associated with different health outcomes between countries.

The results across all models were nearly identical when obtaining marginal effects at the mean.

### **Supplementary Methods S4: Non-Response Weight Derivation Method in BCS70**

***Supplementary Methods Table S4i.*** *Variables used in weight derivation at each age in BCS70.*

| **Variable used to derive weight** | **Weight Age 46** | | **Weight Age 42** | | **Weight Age 34** | |
| --- | --- | --- | --- | --- | --- | --- |
|  | **Age Taken** | **Variable name** | **Age Taken** | **Variable name** | **Age Taken** | **Variable name** |
| *Sex* | Birth | sex | Birth | sex | Birth | sex |
| *Parental Social Class* | Birth | BD1PSOC | Birth | BD1PSOC | Birth | BD1PSOC |
| *Number of rooms in house* | 5 | e228a | 5 | e228a | 5 | e228a |
| *Cognitive ability* | 10 | cog_10 * | 10 | cog_10* | 10 | cog_10 * |
| *Malaise Score* | 16 | BD4MAL | 16 | BD4MAL | 16 | BD4MAL |
| *Voting* | 42 | B9SCQ6 | 34 | b7vote01 | 29 | vote97 |
| *Membership in social/political/sport organisations* | 42 | org_42 * | 34 | org_34 * | 29 | org_29 * |
| *Educational Qualification* | 42 | BD9HNVQ | 38 | BD8HNVQ | 29 | HINVQ00 |
| *Economic Activity (Whether in employment)* | 42 | BD9ECACT | 38 | b8Econ02 | 29 | econact |
| *Partnership Status (Whether currently or previously married)* | 42 | BD9MS | 38 | b8ms | 29 | marstat2 |
| *Psychological distress (Malaise score)* | 42 | BD9MAL | 34 | BD7MAL | 29 | BD6MAL |
| *BMI* | 42 | BD9BMI | 34 | bd7bmi | 29 | bmi29 * |
| *Self-rated Health* | 42 | B9HLTHGN | 38 | b8hlthgn | 29 | hlthgen |
| *Smoking Status* | 42 | B9SMOKIG | 38 | bd8smoke | 29 | smoking |
| *Social Capital/support (how frequently meets family or friends)* | 42 | B09FAMFREMT * | 34 | bd7vfrnd | 29 | outalone |
| *Social Capital/support (whether people around would be willing to listen to problems)* | 42 | B9LISTEN | Not Available |  | Not Available | - |
| *Income* | 42 | hh_inc42*  hh_inc29 * | 38 | hh_inc38 * | 29 | hh_inc29 * |
|  | 29 |  |  |  |  |  |
| *Indicator of non-response in all previous sweeps* | 42 | OUTCME0_nr_cum42 | 38 | OUTCME0_nr_cum38 | 29 | OUTCME0_nr_cum29 |

***Supplementary Methods Table S4i. Footnote.*** ** Derived from other variables in the dataset. Please see table below.*

***Supplementary Methods Table S4ii.*** *Variables and methods used to create derived variables for weights.*

| **Derived Variable** | **Variables Derived From** | **Method** |
| --- | --- | --- |
| cog_10 | “BCS10simG1” to “BCS10simG21”, “BCS10word1” to “BCS10word37”, “BCS10digit1” to “BCS10digit34”, “BCS10mat1” to “BCS10mat28” ,“PLCT1” to “PLCT103”, “i4101” to “i4164”, BD3MATHS | Principal component analysis. |
| org_42 | “B9SCQ8A” to “B9SCQ8P” | Involvement in different types of organisations was combined across the variables, to indicate numbers of organisations involved in (0, 1, 2 or more). |
| org_34 | “b7fintr1” to “b7fintr7” | Involvement in different types of organisations was combined across the variables, to indicate numbers of organisations involved in (0, 1, 2 or more). |
| org_29 | “orgever1” to “orgever7” | Involvement in different types of organisations was combined across the variables, to indicate numbers of organisations involved in (0, 1, 2 or more). |
| bmi29 | wtstone2, wtpound2, wtkilos2, htmetre2, htcms2, htfeet2, htinche2 | Where necessary, weight was converted to kg, height was converted to meters, and from this BMI was calculated (BMI = weight (kg)/ height(m)^2^ |
| B09FAMFREMT | B9FREMT, B9FAMTDR, B9FAMMT | Frequency of meeting friends or family was combined across variables (never/rarely, fairly frequently, very frequently). |
| hh_inc42 | Multiple income and employment related variables at age 42. | Full method to create standardised household income will be published when deposited with UK Data Service. |
| hh_inc38 | Multiple income and employment related variables at age 38. | Full method to create standardised household income will be published when deposited with UK Data Service. |
| hh_inc29 | Multiple income and employment related variables at age 29. | Full method to create standardised household income will be published when deposited with UK Data Service. |

Weight Derivation Method

Weights were derived for the target population at each sweep, which was those cohort members who were alive and living in the UK at the point of data collection.

Multiple imputation was used to generate weights, to ensure that it was possible to include all cohort members in the weight-derivation process, by imputing the variables indicated in Methods Table S3 (excluding the indicators of non-response in all previous sweeps, as well as response to the current sweep, which were complete variables). Multiple imputation was run under the assumption of missing at random, obtaining 5 imputations each, and combined using Rubin’s rule. Multiple imputation was done separately for the weight at each age.

Following imputation, logistic regression was used to predict probability of responding to the present sweep, using the variables indicated in supplementary methods S4 Table 4i and Table 4ii. Using the regression output, probability of responding was predicted, and inverse probability weights were generated.

Weights at each age were then truncated to the value of 10 in order to prevent extreme weights exerting undue influence. The weights were then rescaled to the respective sweeps.

### **Supplementary Methods S5: Methods used to apply complex survey design across studies.**

Add Health uses a complex, stratified sampling strategy that accounts for the region, urbanicity, size, type, and racial composition of schools from which students were recruited, thus maintaining the national representativeness of the data. The Add Health data set therefore includes a primary sampling unit (PSU) value for each participant, and a strata variable. Add Health also includes survey weights that account for non-representativeness among adults providing biomarker samples.

To allow inclusion of the complex survey design in Add Health when analysing data across the two data sets, dummy PSU and strata were created in BCS70. In BCS70, a unique primary sampling unit (PSU) value was assigned to each cohort member that differed to those used in Add Health, a constant strata value was created across the cohort that also differed to those used in Add Health therefore allowing use of the complex survey characteristics in Add Health in the pooled analysis.

### **Supplementary Results S1: Unweighted distribution of covariates and outcomes**

***Supplementary Results Table S1.*** *Unweighted distribution of covariates and outcomes in BCS70 and Add Health (non-Hispanic White only; among participants with valid weights)*

|  | **BCS70** | **Add Health** |
| --- | --- | --- |
|  | ***N*** | ***N*** |
| ***Alcohol Consumption*** |  |  |
| Heavy Drinker | 1958 | 807 |
| Non-Heavy Drinker | 7697 | 5863 |
| Total | 9655 | 6670 |
| ***Smoking Status*** |  |  |
| Regular Smoker | 2373 | 1255 |
| Non-Smoker | 7261 | 5422 |
| Total | 9634 | 6677 |
| ***Self-Rated Health*** |  |  |
| Poor/Fair Health | 1488 | 771 |
| Good/Excellent Health | 8310 | 5948 |
| Total | 9798 | 6719 |
| ***Obesity*** |  |  |
| Obese | 2873 | 2644 |
| Not Obese | 5621 | 4047 |
| Total | 8494 | 6691 |
| ***Blood Pressure (Biomarker)*** |  |  |
| Hypertension | 1418 | 668 |
| Normal | 6111 | 2465 |
| Total | 7529 | 3133 |
| ***Cholesterol (Biomarker)*** |  |  |
| Unhealthy | 423 | 269 |
| Healthy | 5613 | 2556 |
| Total | 6036 | 2825 |
| ***HbA1C (Biomarker)*** |  |  |
| Diabetes | 323 | 124 |
| No Diabetes | 5673 | 2662 |
| Total | 5996 | 2786 |
| ***Any Hypertension*** |  |  |
| Yes | 1435 | 992 |
| No | 6111 | 2465 |
| Total | 7546 | 3457 |
| ***Any High Cholesterol*** |  |  |
| Yes | 514 | 420 |
| No | 5613 | 2556 |
| Total | 6127 | 2976 |
| ***Any Diabetes*** |  |  |
| Yes | 384 | 219 |
| No | 5673 | 2662 |
| Total | 6057 | 2881 |
| ***Sex*** |  |  |
| Male | 4272 | 3020 |
| Female | 4688 | 3707 |
| Total | 8960 | 6727 |
| ***Parental Education Level*** |  |  |
| Neither parent has a degree | 3808 | 4099 |
| At least one parent has a degree | 1043 | 2551 |
| Total | 4851 | 6650 |
| ***Own Education Level  (Add Health Wave V,  BCS70 Sweep 9)*** |  |  |
| No university degree | 5773 | 3787 |
| Degree-level educated | 4060 | 2932 |
| Total | 9833 | 6715 |
| ***Own Education Level  (BCS70 Sweep 8 only)*** |  |  |
| No university degree | 5882 | - |
| Degree-level educated | 3783 | - |
| Total | 9665 | - |
| ***Own Income  (Add Health Wave V,  BCS70 Sweep 9)*** |  |  |
| Lowest income quintile | 1582 | 902 |
| Second quintile | 1589 | 1348 |
| Middle quintile | 1581 | 1030 |
| Fourth quintile | 1581 | 1333 |
| Highest income quintile | 1575 | 1126 |
| Total | 7908 | 5739 |
| ***Own Income  (BCS70 Sweep 8 only)*** |  |  |
| Lowest income quintile | 1831 | - |
| Second quintile | 1831 | - |
| Middle quintile | 1822 | - |
| Fourth quintile | 1820 | - |
| Highest income quintile | 1822 | - |
| Total | 9126 | - |

### **Supplementary Results S2: Results table showing marginal estimates from modified Poisson regression for Model 2**

***Supplementary Results Table S2.*** *Marginal estimates from modified Poisson regression for Model 2, examining country differences in SEP inequalities health outcomes.*

|  |  | **Britain** | | | **United States** | | |  |
| --- | --- | --- | --- | --- | --- | --- | --- | --- |
| **Health Outcome** | **Level of SEP** | **Estimates** | **Lower 95% CI** | **Upper 95% CI** | **Estimates** | **Lower 95% CI** | **Upper 95% CI** | **P Value Difference ^D^** |
| Heavy Drinking | Parents ^A^ = no degree | 0·195 | 0·181 | 0·208 | 0·112 | 0·099 | 0·126 | 0·64 |
|  | Parents ^A^ = degree | 0·251 | 0·223 | 0·279 | 0·133 | 0·117 | 0·150 |  |
|  | Own ^B^ = no degree | 0·187 | 0·176 | 0·199 | 0·125 | 0·110 | 0·140 | 0·44 |
|  | Own ^B^ = degree | 0·203 | 0·189 | 0·217 | 0·113 | 0·096 | 0·130 |  |
|  | Income ^C^ = Bottom | 0·168 | 0·148 | 0·188 | 0·123 | 0·096 | 0·151 | 0·68 |
|  | Income ^C^ = Middle | 0·192 | 0·171 | 0·214 | 0·110 | 0·086 | 0·135 | 0·21 |
|  | Income ^C^ = Top | 0·234 | 0·211 | 0·256 | 0·121 | 0·096 | 0·146 | 0·022 |
| Smoking | Parents ^A^ = no degree | 0·234 | 0·219 | 0·249 | 0·272 | 0·249 | 0·296 | 0·0005 |
|  | Parents ^A^ = degree | 0·141 | 0·118 | 0·164 | 0·110 | 0·094 | 0·126 |  |
|  | Own ^B^ = no degree | 0·336 | 0·321 | 0·351 | 0·317 | 0·295 | 0·340 | <0·0001 |
|  | Own ^B^ = degree | 0·174 | 0·160 | 0·189 | 0·058 | 0·049 | 0·068 |  |
|  | Income ^C^ = Bottom | 0·416 | 0·387 | 0·445 | 0·459 | 0·421 | 0·497 | 0·0002 |
|  | Income ^C^ = Middle | 0·249 | 0·226 | 0·273 | 0·169 | 0·131 | 0·207 | 0·802 |
|  | Income ^C^ = Top | 0·182 | 0·160 | 0·203 | 0·067 | 0·046 | 0·087 | 0·12 |
| Self-rated Health | Parents ^A^ = no degree | 0·147 | 0·134 | 0·160 | 0·143 | 0·126 | 0·161 | 0·307 |
|  | Parents ^A^ = degree | 0·099 | 0·077 | 0·120 | 0·077 | 0·060 | 0·094 |  |
|  | Own ^B^ = no degree | 0·222 | 0·207 | 0·237 | 0·172 | 0·153 | 0·190 | 0·29 |
|  | Own ^B^ = degree | 0·110 | 0·098 | 0·123 | 0·044 | 0·035 | 0·054 |  |
|  | Income ^C^ = Bottom | 0·334 | 0·300 | 0·369 | 0·270 | 0·230 | 0·309 | 0·11 |
|  | Income ^C^ = Middle | 0·133 | 0·111 | 0·155 | 0·094 | 0·068 | 0·120 | 0·0030 |
|  | Income ^C^ = Top | 0·062 | 0·048 | 0·076 | 0·029 | 0·015 | 0·043 | 0·0002 |
| Obesity | Parents ^A^ = no degree | 0·351 | 0·332 | 0·370 | 0·445 | 0·421 | 0·468 | 0·72 |
|  | Parents ^A^ = degree | 0·251 | 0·221 | 0·281 | 0·334 | 0·300 | 0·368 |  |
|  | Own ^B^ = no degree | 0·372 | 0·354 | 0·391 | 0·465 | 0·444 | 0·486 | 0·0003 |
|  | Own ^B^ = degree | 0·300 | 0·283 | 0·317 | 0·312 | 0·282 | 0·342 |  |
|  | Income ^C^ = Bottom | 0·358 | 0·320 | 0·396 | 0·501 | 0·454 | 0·549 | 0·34 |
|  | Income ^C^ = Middle | 0·366 | 0·334 | 0·397 | 0·425 | 0·390 | 0·459 | 0·203 |
|  | Income ^C^ = Top | 0·253 | 0·228 | 0·279 | 0·236 | 0·199 | 0·273 | 0·0003 |
| High BP (Biomarker) | Parents ^A^ = no degree | 0·190 | 0·174 | 0·205 | 0·238 | 0·208 | 0·268 | 0·95 |
|  | Parents ^A^ = degree | 0·156 | 0·131 | 0·181 | 0·203 | 0·178 | 0·227 |  |
|  | Own ^B^ = no degree | 0·199 | 0·184 | 0·214 | 0·260 | 0·228 | 0·292 | 0·095 |
|  | Own ^B^ = degree | 0·162 | 0·147 | 0·177 | 0·183 | 0·158 | 0·207 |  |
|  | Income ^C^ = Bottom | 0·189 | 0·157 | 0·222 | 0·266 | 0·206 | 0·326 | 0·4000 |
|  | Income ^C^ = Middle | 0·204 | 0·177 | 0·230 | 0·215 | 0·167 | 0·263 | 0·53 |
|  | Income ^C^ = Top | 0·161 | 0·137 | 0·185 | 0·166 | 0·130 | 0·203 | 0·402 |
| High Cholesterol (Biomarker) | Parents ^A^ = no degree | 0·064 | 0·053 | 0·075 | 0·126 | 0·103 | 0·149 | 0·085 |
|  | Parents ^A^ = degree | 0·060 | 0·037 | 0·082 | 0·083 | 0·060 | 0·107 |  |
|  | Own ^B^ = no degree | 0·086 | 0·072 | 0·099 | 0·137 | 0·112 | 0·162 | 0·037 |
|  | Own ^B^ = degree | 0·059 | 0·048 | 0·069 | 0·070 | 0·051 | 0·089 |  |
|  | Income ^C^ = Bottom | 0·098 | 0·066 | 0·131 | 0·181 | 0·122 | 0·240 | 0·52 |
|  | Income ^C^ = Middle | 0·081 | 0·060 | 0·102 | 0·072 | 0·042 | 0·103 | 0·060 |
|  | Income ^C^ = Top | 0·051 | 0·035 | 0·068 | 0·076 | 0·036 | 0·116 | 0·33 |
| High HbA1c (Biomarker) | Parents ^A^ = no degree | 0·060 | 0·049 | 0·071 | 0·056 | 0·042 | 0·070 | 0·804 |
|  | Parents ^A^ = degree | 0·026 | 0·013 | 0·040 | 0·026 | 0·015 | 0·036 |  |
|  | Own ^B^ = no degree | 0·073 | 0·060 | 0·086 | 0·055 | 0·041 | 0·069 | 0·54 |
|  | Own ^B^ = degree | 0·041 | 0·032 | 0·050 | 0·031 | 0·017 | 0·045 |  |
|  | Income ^C^ = Bottom | 0·069 | 0·045 | 0·093 | 0·079 | 0·046 | 0·112 | 0·25 |
|  | Income ^C^ = Middle | 0·067 | 0·045 | 0·089 | 0·048 | 0·020 | 0·077 | 0·99 |
|  | Income ^C^ = Top | 0·046 | 0·030 | 0·063 | 0·010 | -0·001 | 0·021 | 0·35 |
| Any Hypertension | Parents ^A^ = no degree | 0·191 | 0·175 | 0·207 | 0·328 | 0·294 | 0·362 | 0·47 |
|  | Parents ^A^ = degree | 0·158 | 0·133 | 0·183 | 0·275 | 0·245 | 0·306 |  |
|  | Own ^B^ = no degree | 0·201 | 0·186 | 0·217 | 0·359 | 0·326 | 0·392 | 0·0012 |
|  | Own ^B^ = degree | 0·164 | 0·149 | 0·179 | 0·240 | 0·209 | 0·272 |  |
|  | Income ^C^ = Bottom | 0·189 | 0·157 | 0·221 | 0·351 | 0·286 | 0·416 | 0·47 |
|  | Income ^C^ = Middle | 0·205 | 0·178 | 0·232 | 0·304 | 0·250 | 0·359 | 0·54 |
|  | Income ^C^ = Top | 0·164 | 0·140 | 0·187 | 0·219 | 0·176 | 0·263 | 0·047 |
| Any High Cholesterol | Parents ^A^ = no degree | 0·077 | 0·064 | 0·089 | 0·187 | 0·159 | 0·214 | 0·016 |
|  | Parents ^A^ = degree | 0·070 | 0·046 | 0·093 | 0·118 | 0·087 | 0·150 |  |
|  | Own ^B^ = no degree | 0·105 | 0·090 | 0·120 | 0·201 | 0·170 | 0·232 | 0·0046 |
|  | Own ^B^ = degree | 0·069 | 0·058 | 0·080 | 0·105 | 0·082 | 0·128 |  |
|  | Income ^C^ = Bottom | 0·129 | 0·094 | 0·163 | 0·266 | 0·206 | 0·327 | 0·087 |
|  | Income ^C^ = Middle | 0·091 | 0·070 | 0·113 | 0·113 | 0·069 | 0·158 | 0·35 |
|  | Income ^C^ = Top | 0·057 | 0·040 | 0·074 | 0·101 | 0·054 | 0·147 | 0·66 |
| Any Diabetes | Parents ^A^ = no degree | 0·069 | 0·056 | 0·081 | 0·107 | 0·085 | 0·129 | 0·083 |
|  | Parents ^A^ = degree | 0·033 | 0·018 | 0·047 | 0·044 | 0·030 | 0·057 |  |
|  | Own ^B^ = no degree | 0·086 | 0·071 | 0·100 | 0·113 | 0·089 | 0·136 | 0·077 |
|  | Own ^B^ = degree | 0·049 | 0·040 | 0·059 | 0·047 | 0·031 | 0·063 |  |
|  | Income ^C^ = Bottom | 0·105 | 0·075 | 0·135 | 0·166 | 0·116 | 0·217 | 0·070 |
|  | Income ^C^ = Middle | 0·069 | 0·048 | 0·090 | 0·090 | 0·053 | 0·126 | 0·46 |
|  | Income ^C^ = Top | 0·047 | 0·031 | 0·064 | 0·015 | 0·003 | 0·026 | 0·13 |

***Supplementary Results Table S2 Footnote:*** *Results presented are for model 2, exploring country differences in socioeconomic inequalities in health outcomes between the UK and US. ^A^ “Parents” refers to whether either parent has a university bachelor’s degree; ^B^ “own” refers to whether the cohort member has a university bachelor’s degree; ^C^ refers to the cohort members household income; ^D^ P-value Difference refers to whether the magnitude of inequality between the UK and US is statistically different, as determined by Wald-tests. For parental and own education, this is whether the size of the inequality between those with and without a university bachelor’s degree statistically differ between the two countries. For income, the inequality is assessed by how the respective quintile relates to the 20^th^-40^th^ percentile. Therefore, country differences in the inequality are assessed by whether the size of the inequality between the respective income quintile (e.g., top, middle, bottom) and the 20^th^- 40^th^ percentile is statistically different between the two countries. Note: SEP is Socioeconomic Position; BP is Blood pressure; high cholesterol measured by total cholesterol (TC) to High-density lipoprotein (HDL) ratio; HbA1c is Glycated haemoglobin (blood sugar levels). Outcomes labelled “any” refer to biomarkers that have been supplemented with medication use, therefore indicating a positive diagnosis of diseases.*

### **Supplementary Results S3: Sex-stratified analysis - Model 2 in females (figure 3a) and males (figure 3b).**

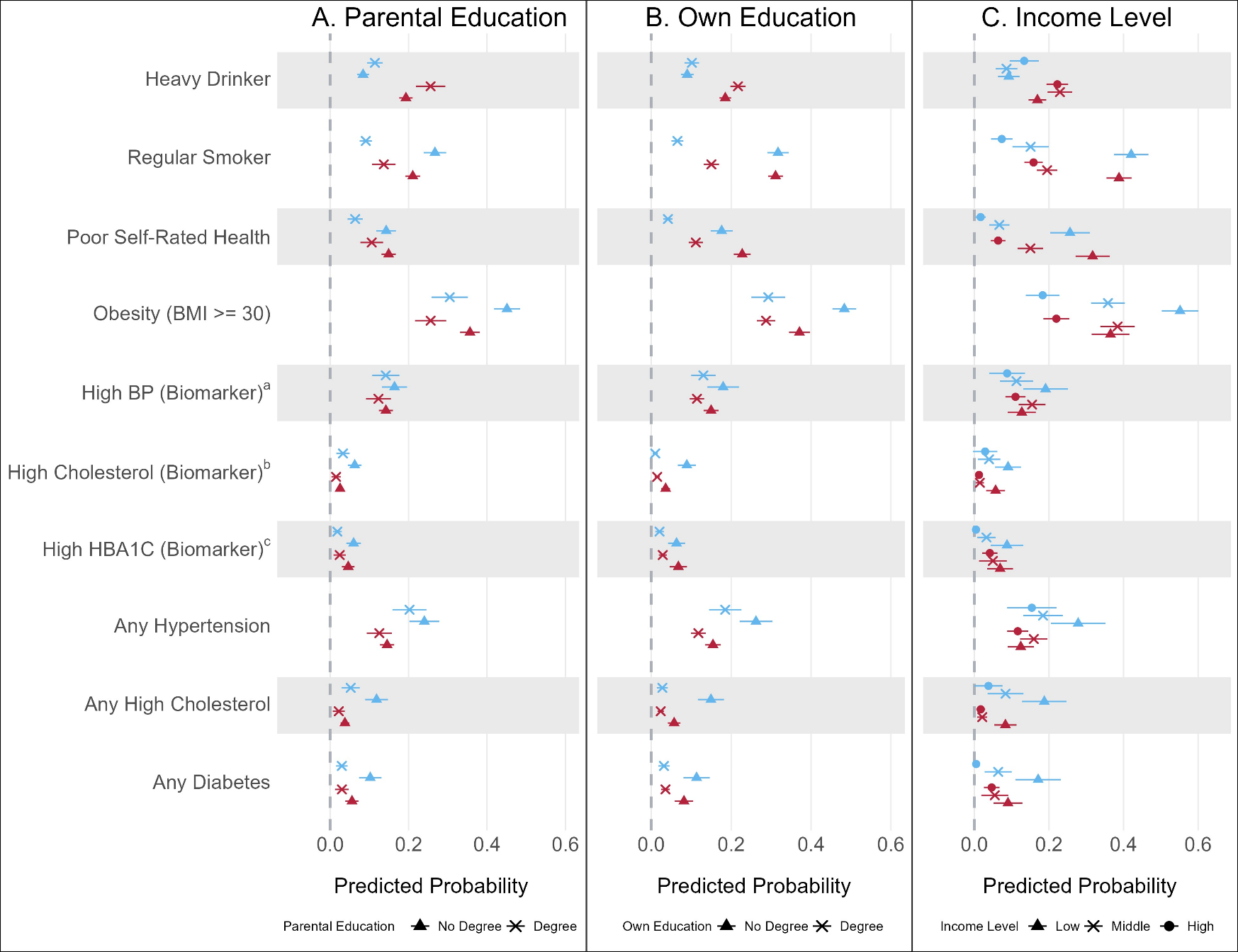
**Supplementary Figure 3a.** Analytic Model 2 in females only: Predicted probabilities from modified Poisson regression showing socioeconomic inequalities in midlife health between Britain and the US (Model 2a, 2b and 2c).

**Supplementary Figure 3b.** Analytic Model 2 in males only: Predicted probabilities from modified Poisson regression showing socioeconomic inequalities in midlife health between Britain and the US (Model 2a, 2b and 2c).

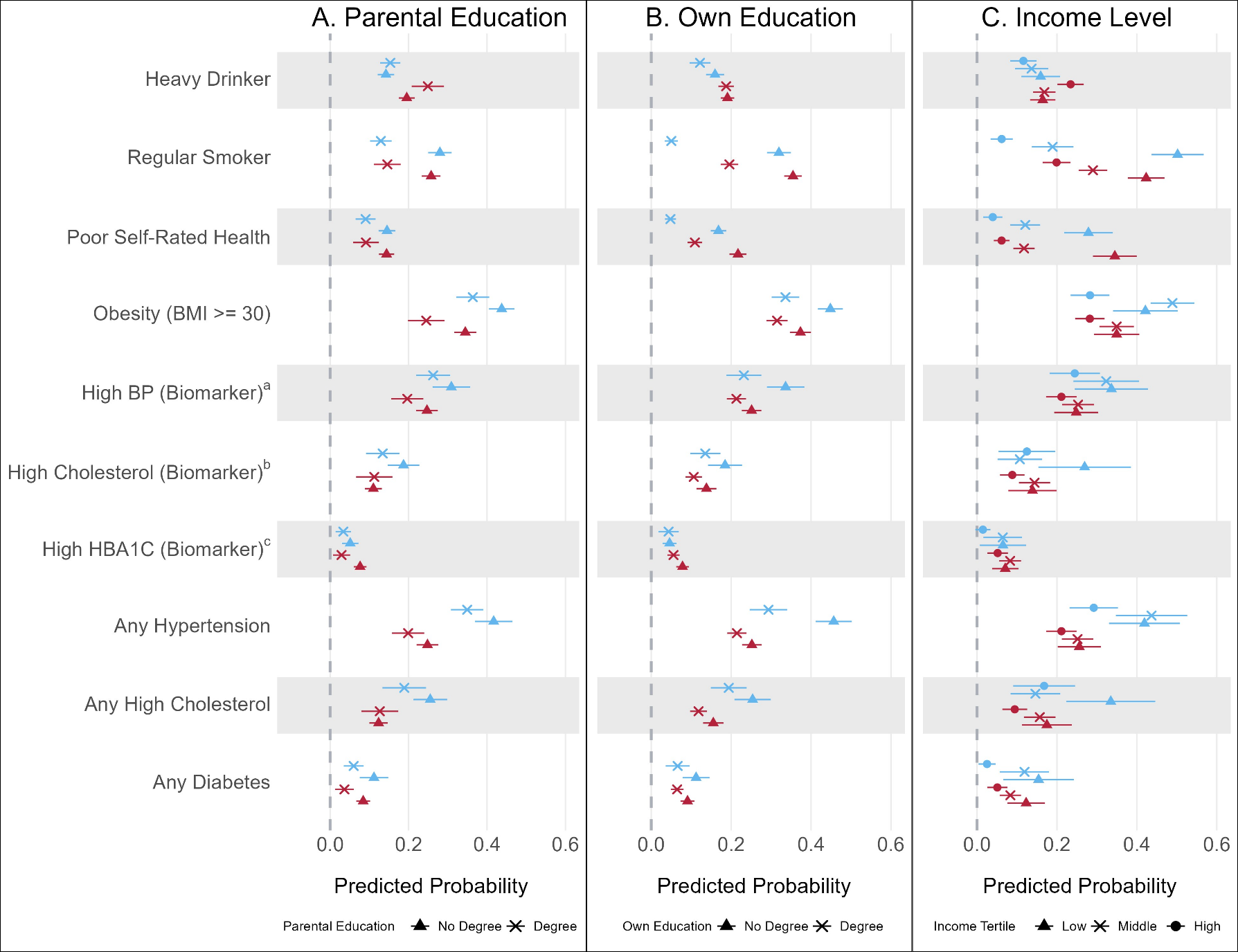

### **Supplementary Results S4: Sex-stratified analysis - Model 3 in females (figure 4a) and males (figure 4b).**

**Supplementary Figure 4a.** Analytic Model 3 in females only: Predicted probabilities from modified Poisson regression of associations with parental education, adjusted for cohort members own SEP (education level and household income) in adulthood.

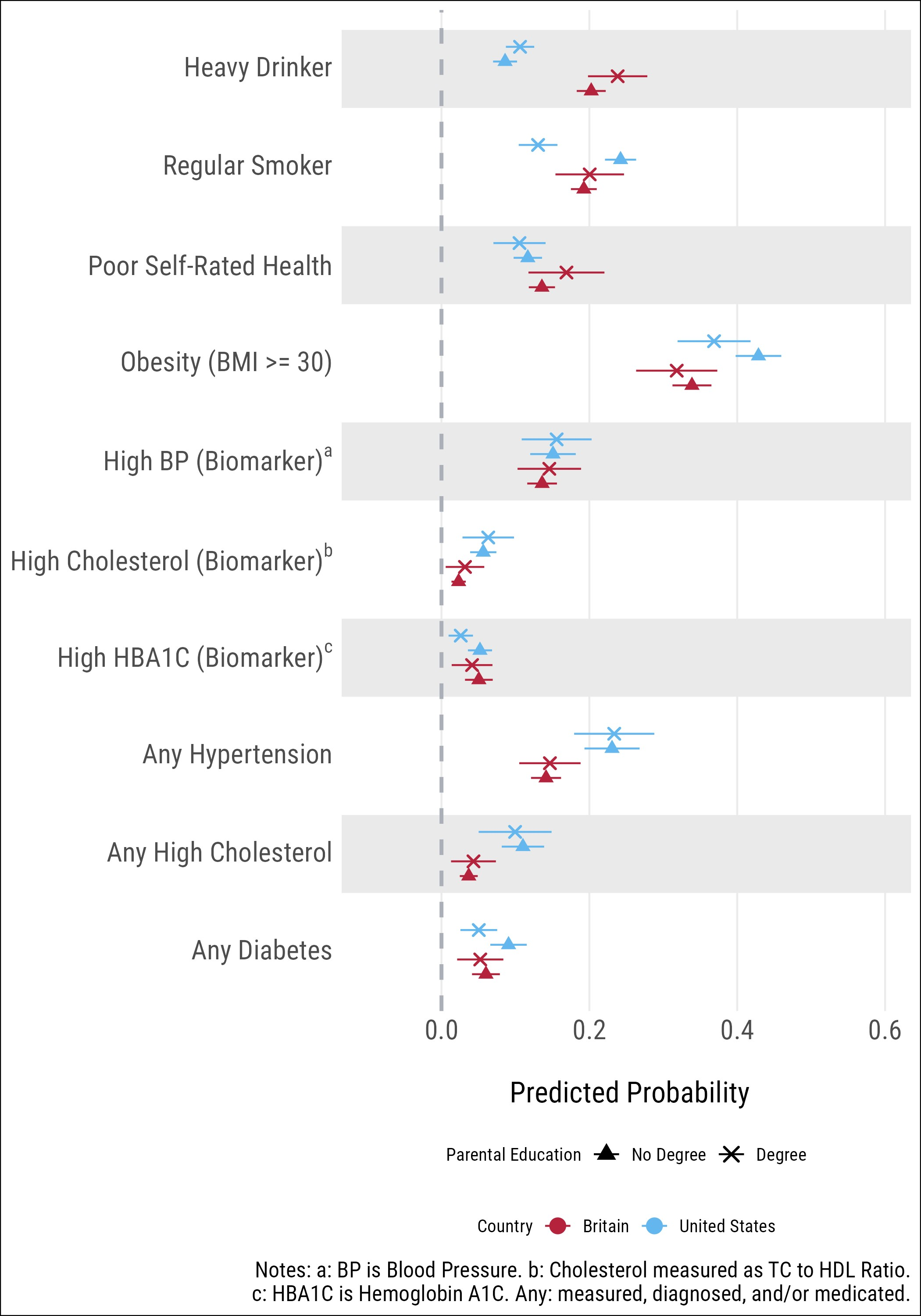

**Supplementary Figure S4b.** Analytic Model 3 in males only: Predicted probabilities from modified Poisson regression of associations with parental education, adjusted for cohort members own SEP (education level and household income) in adulthood.

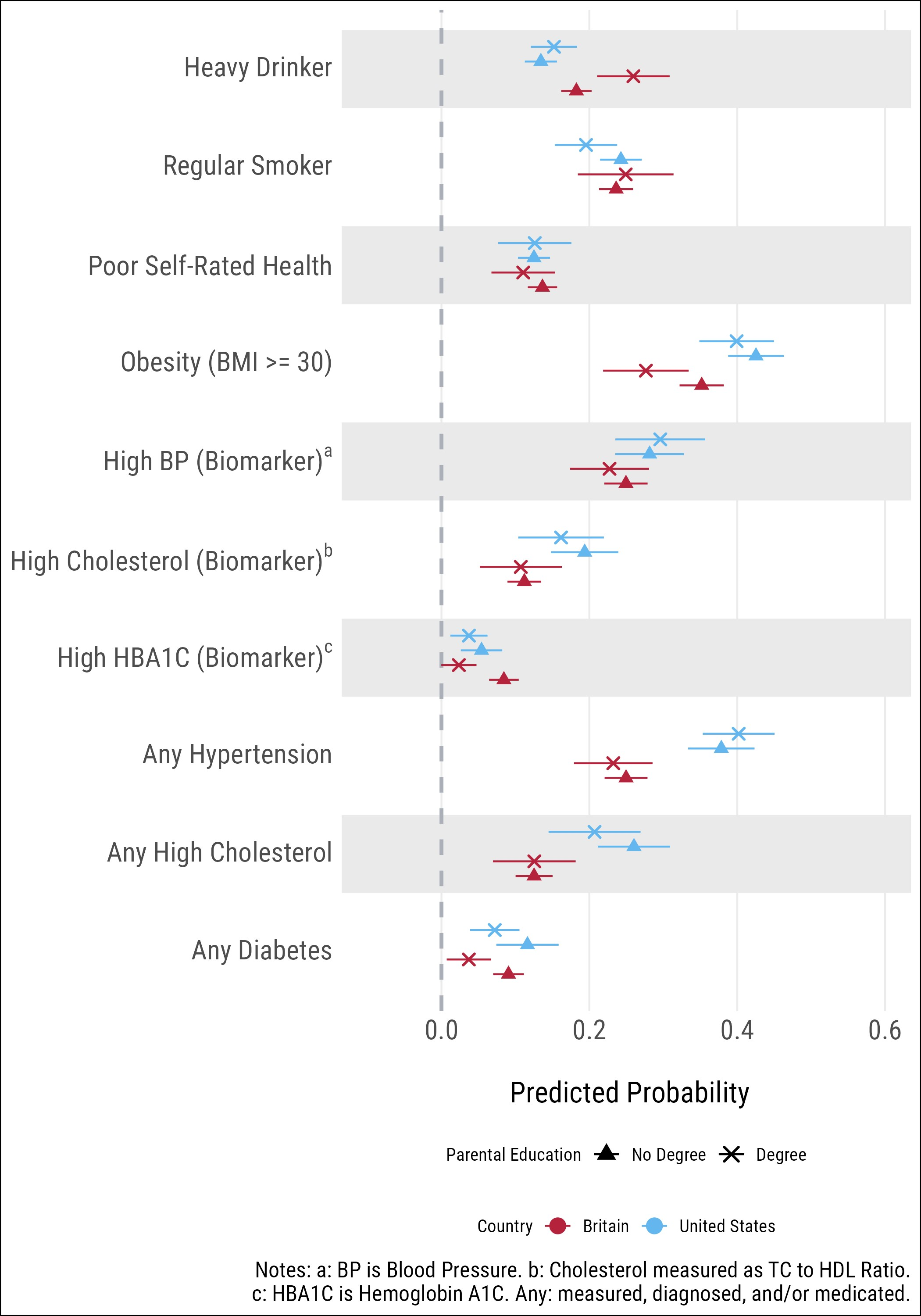

### **Supplementary Results S5: Sensitivity analysis using full, ethnically heterogenous, Add Health Sample**

**Supplementary Figure S5.** Analytic Model 1 using the full ethnically heterogeneous Add Health Sample: Predicted probabilities from modified Poisson regression, comparing health indicators between Britain and the US, by sex (Model 1).

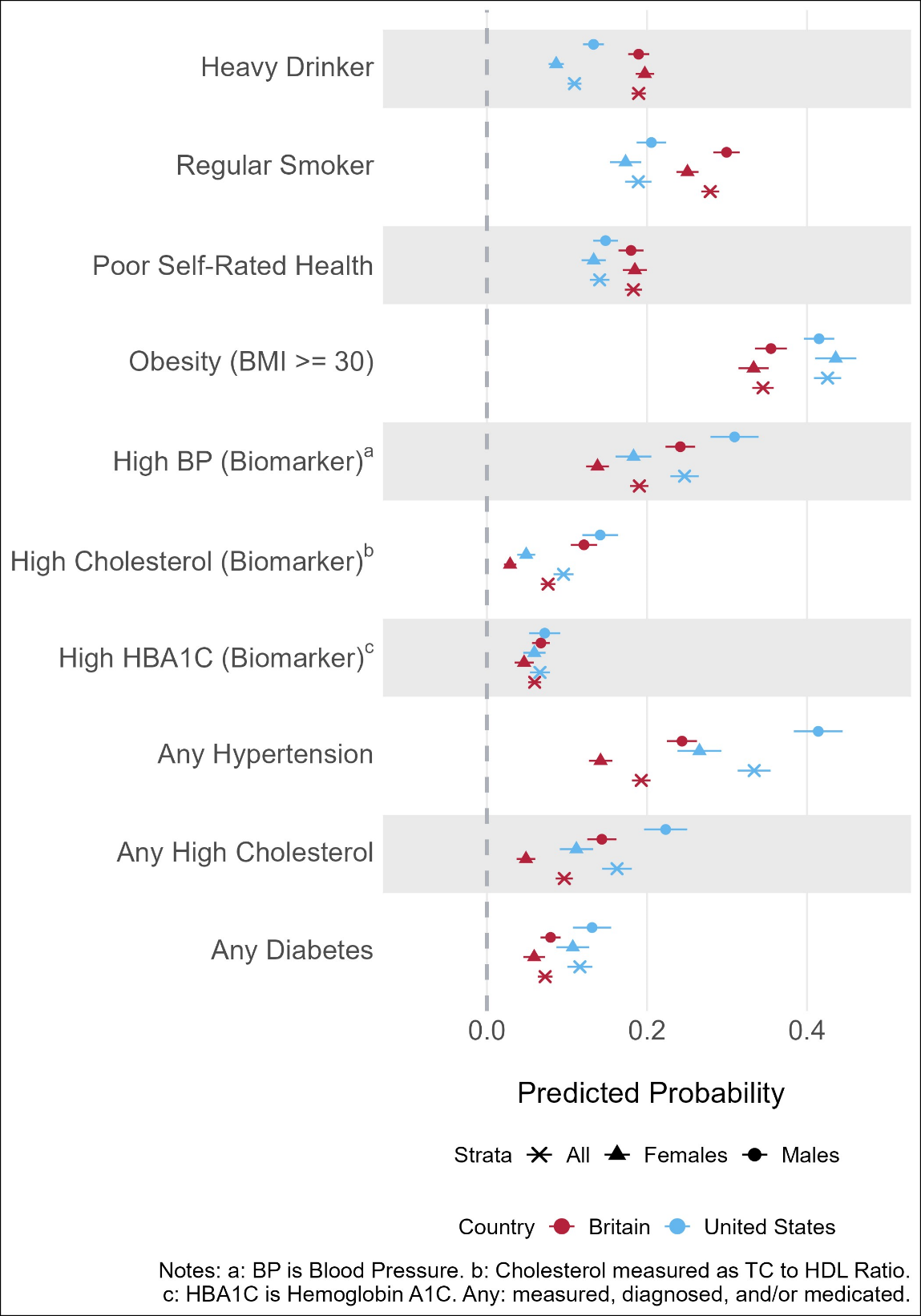

### **Supplementary Results S6: Sensitivity analysis using only directly measured BMI in Add Health**

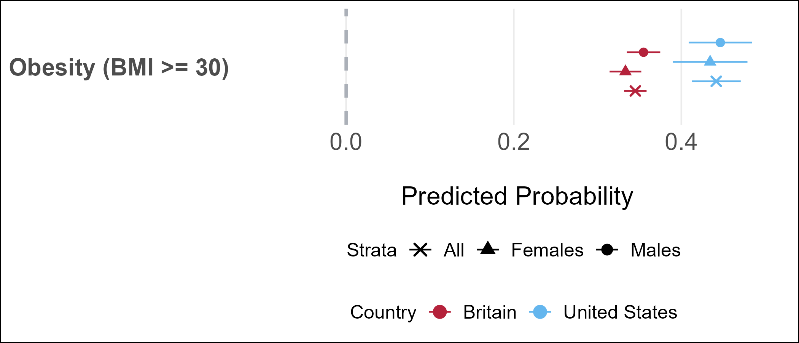
**Supplementary figure 6a.** Analytic model 1, using only directly measured obesity in Add Health.

**Supplementary figure 6b.** Model 2, using only directly measured obesity in Add Health.

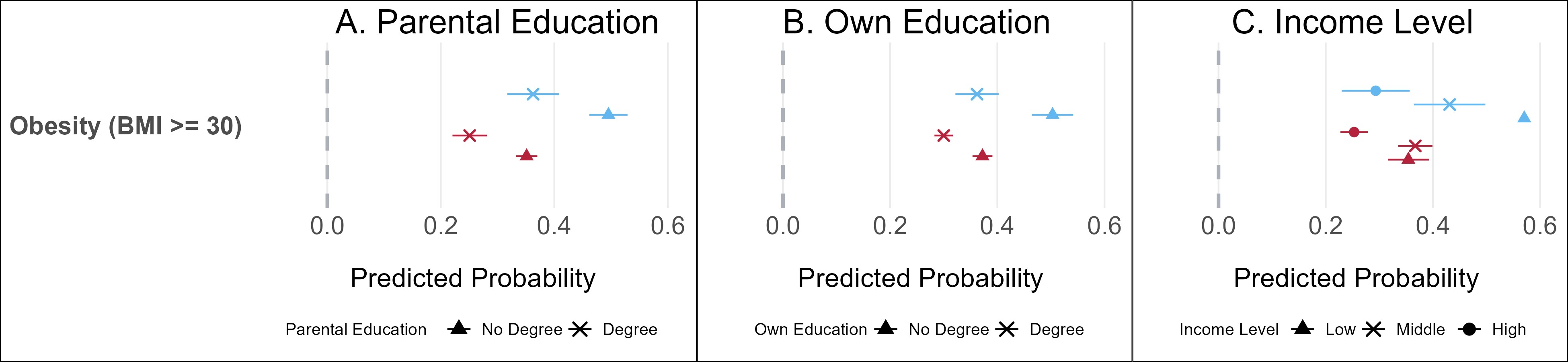

**Supplementary figure 6c.** Model 2 in females, using only directly measured obesity in Add Health.

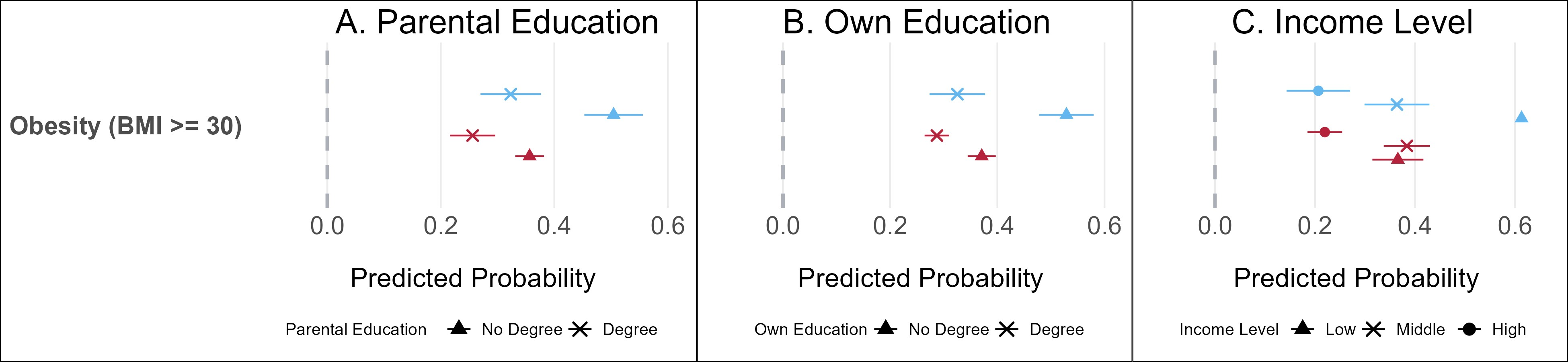

**Supplementary figure 6d.** Model 2 in males, using only directly measured obesity in Add Health.

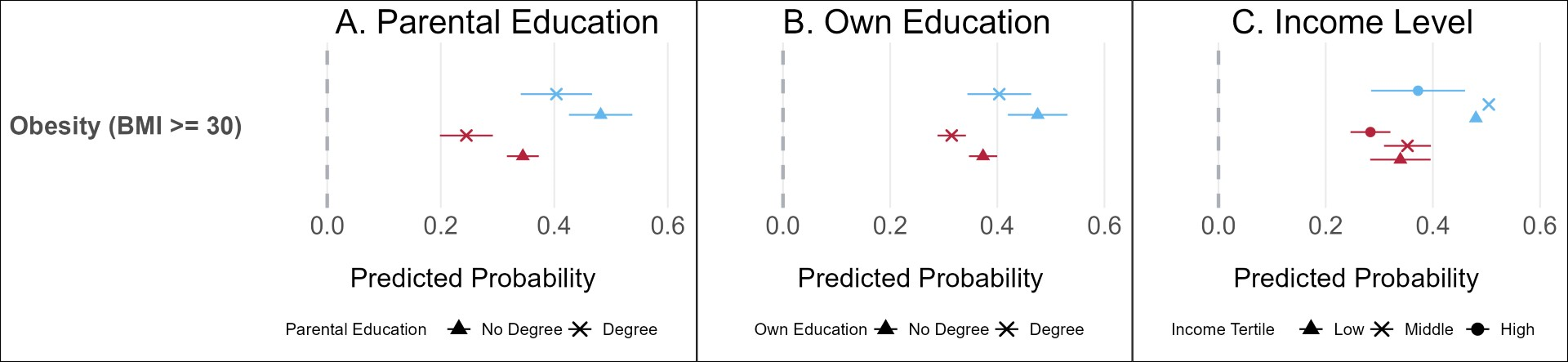

**Supplementary figure 6e.** Analytic Model 3, using only directly measured obesity in Add Health.

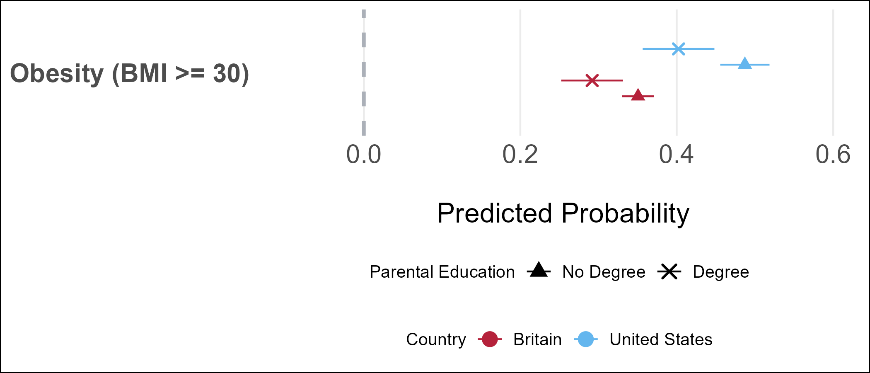

**Supplementary figure 6f.** Model 3 in female, using only directly measured obesity in Add Health.

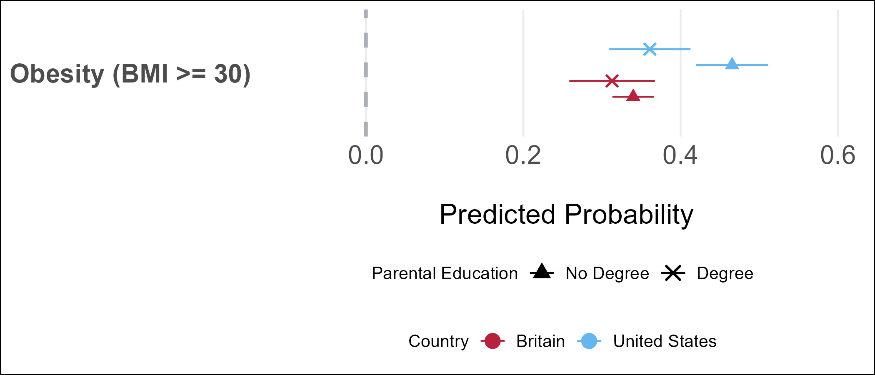

**Supplementary figure 6g.** Model 3 in males, using only directly measured obesity in Add Health.

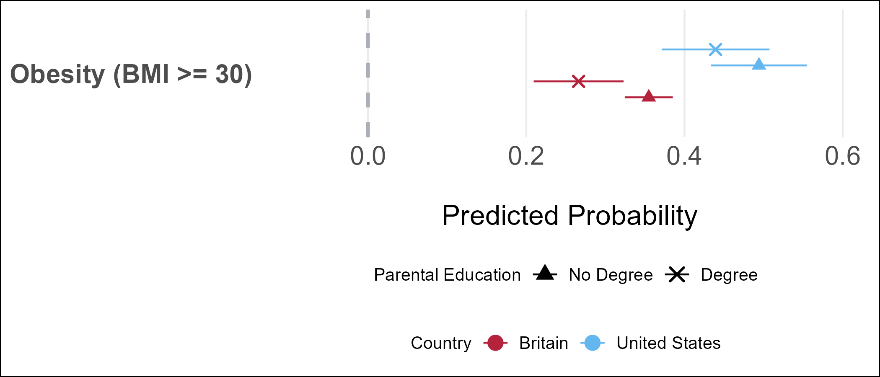

### **Supplementary Results S7: Sensitivity analysis comparing different operationalisations of heavy drinking variable.**

**Supplementary Figure 7a.** Model 1 comparing difference operationalisations of heavy drinking.

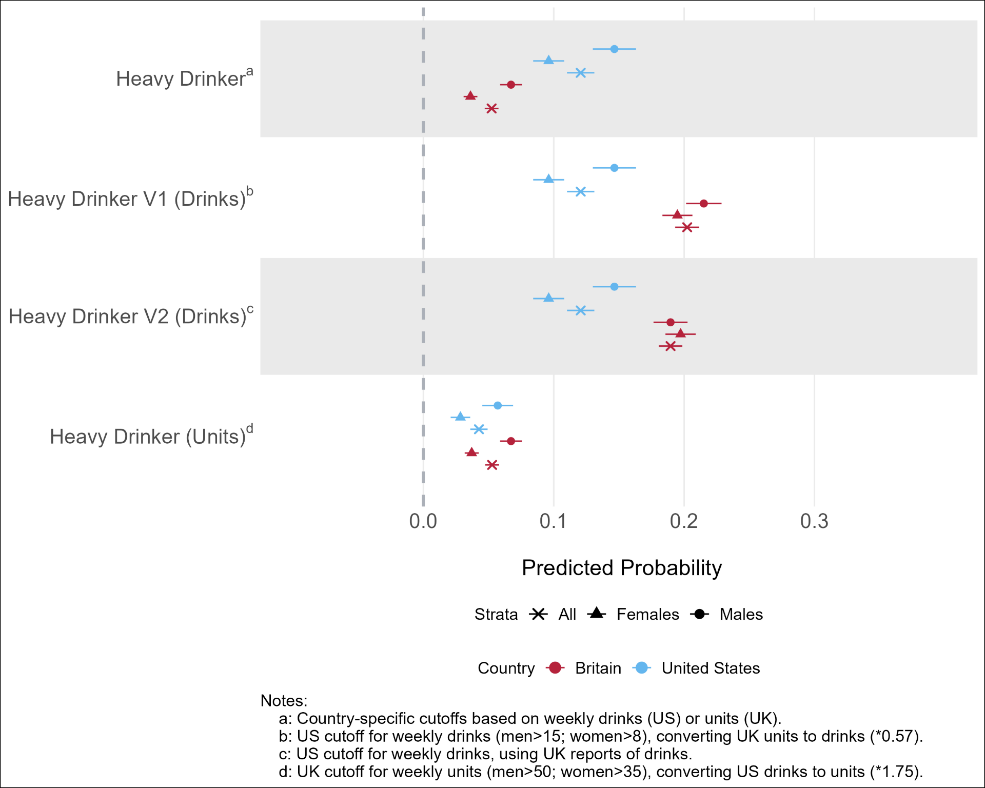

**Supplementary Figure 7b.** Model 2 comparing difference operationalisations of heavy drinking.

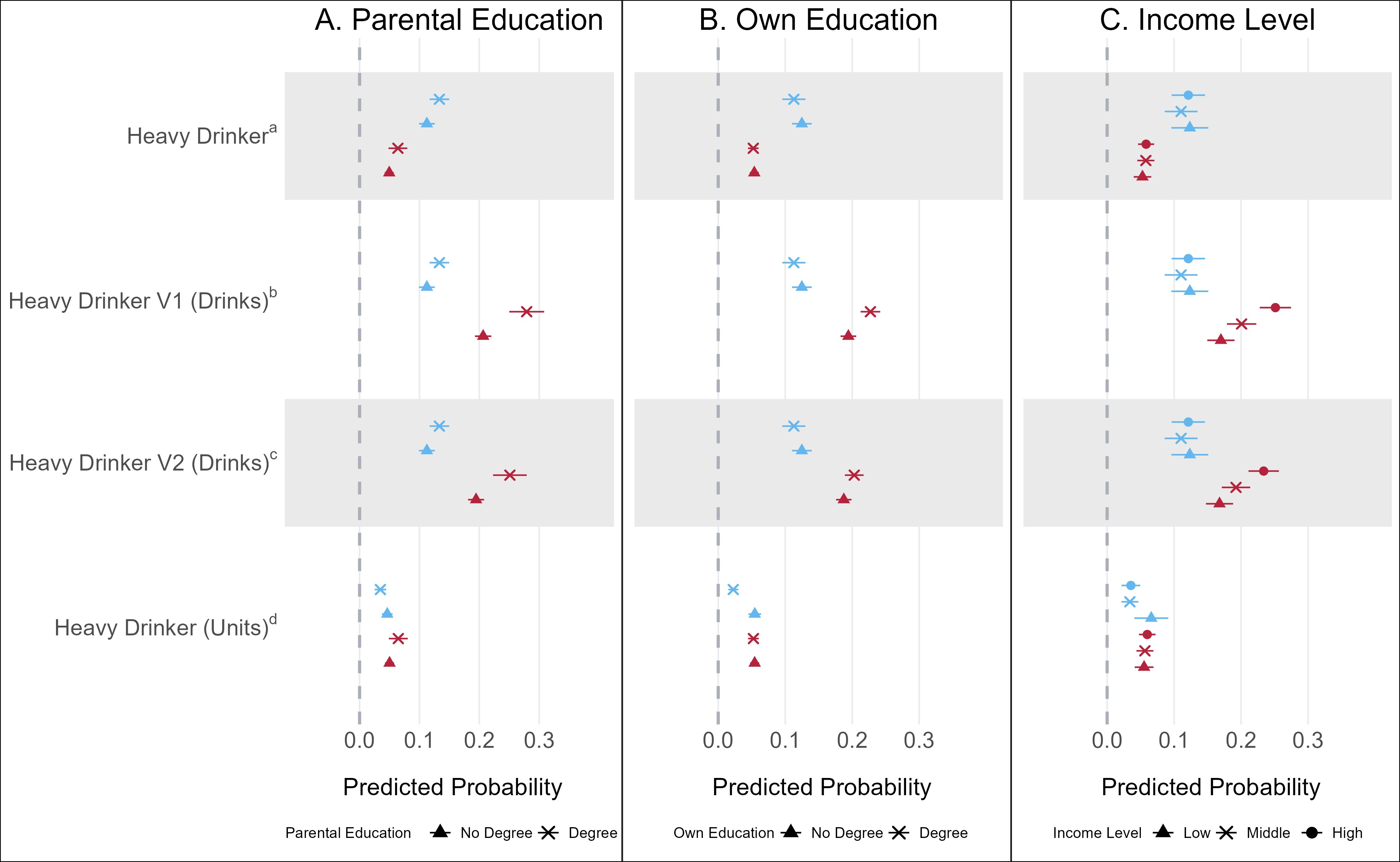

**Supplementary Figure 7c.** Model 2 in females comparing difference operationalisations of heavy drinking.

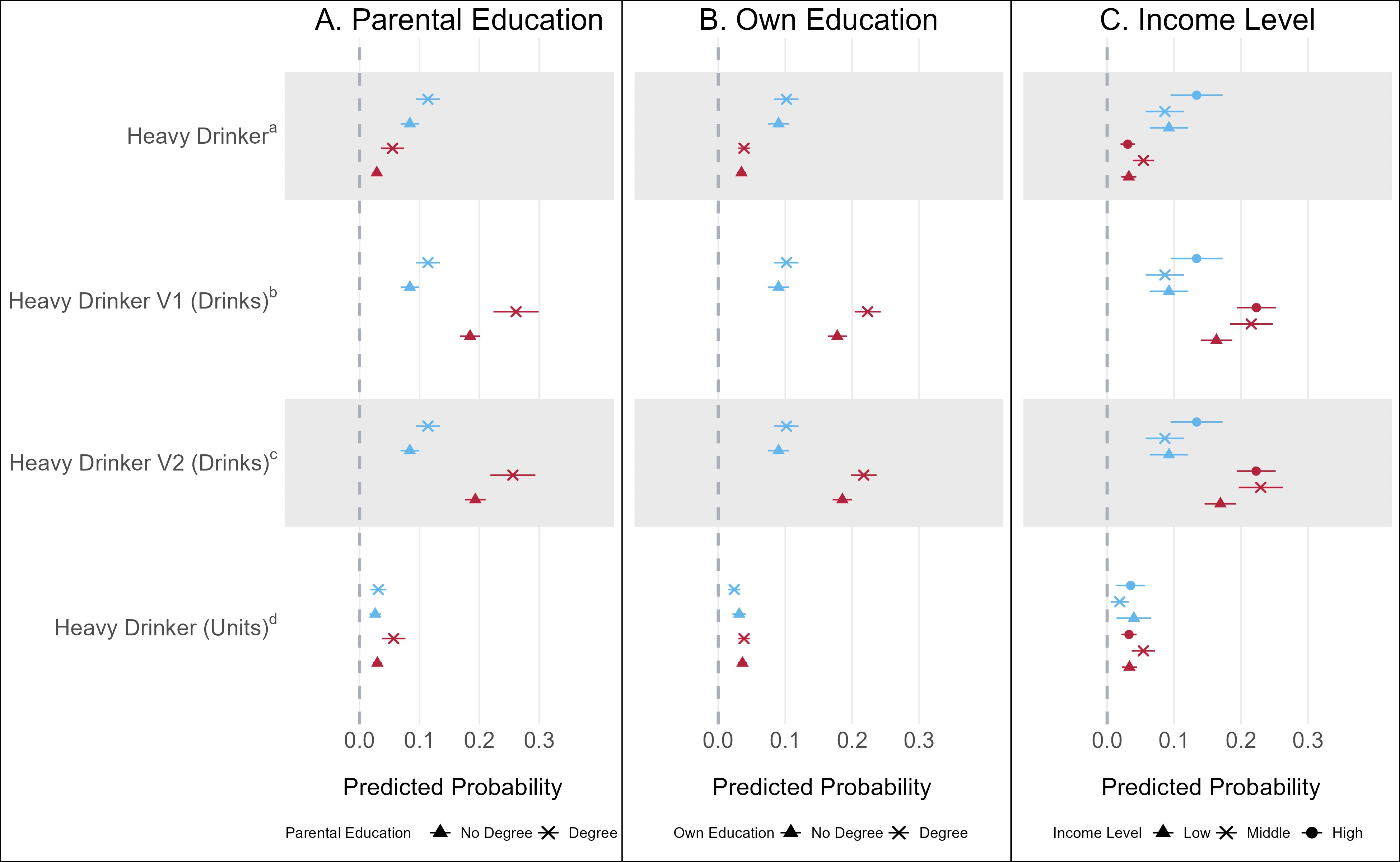

**Supplementary Figure 7d.** Model 2 in males comparing difference operationalisations of heavy drinking.

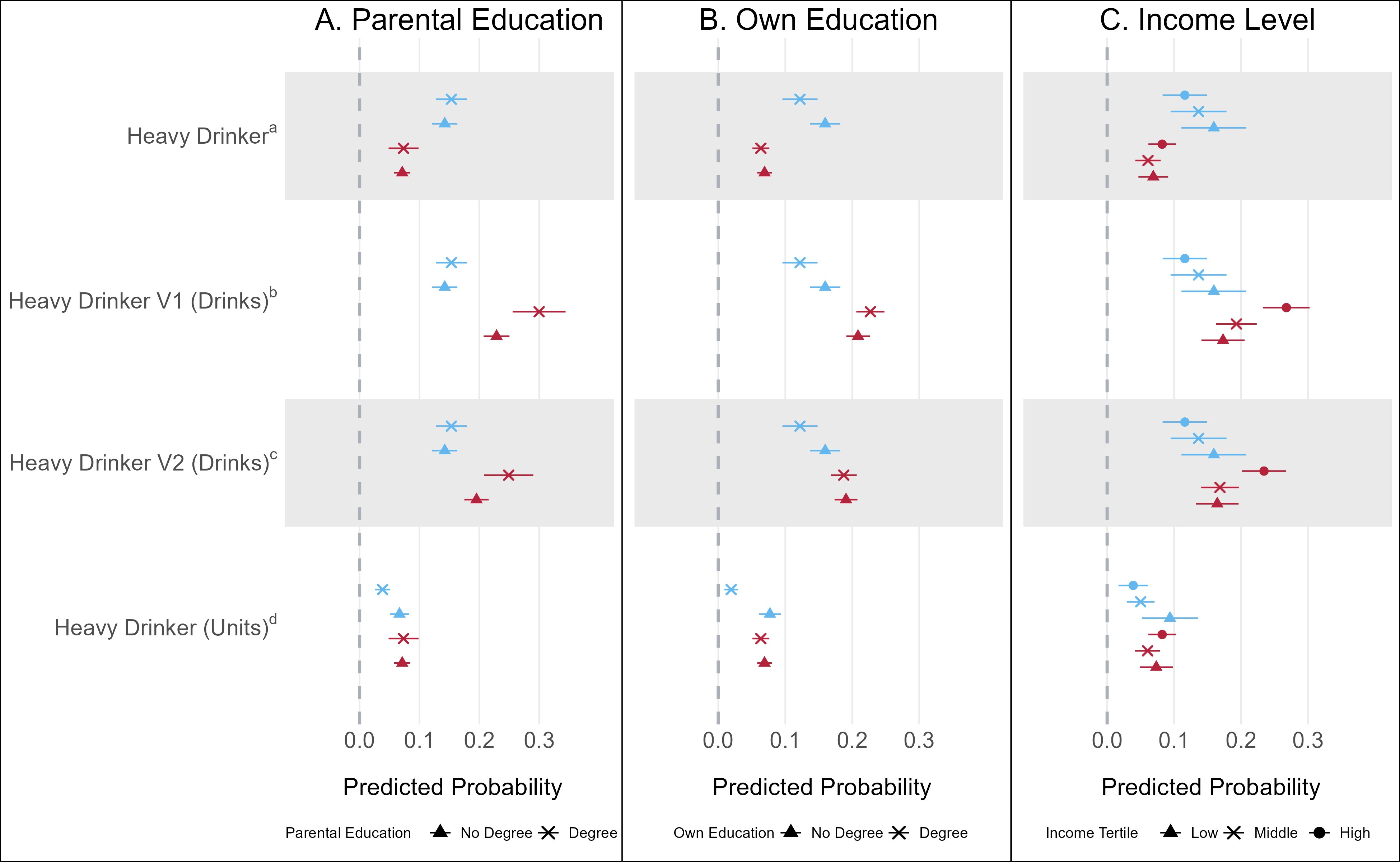

**Supplementary Figure 7e.** Model 3 comparing difference operationalisations of heavy drinking

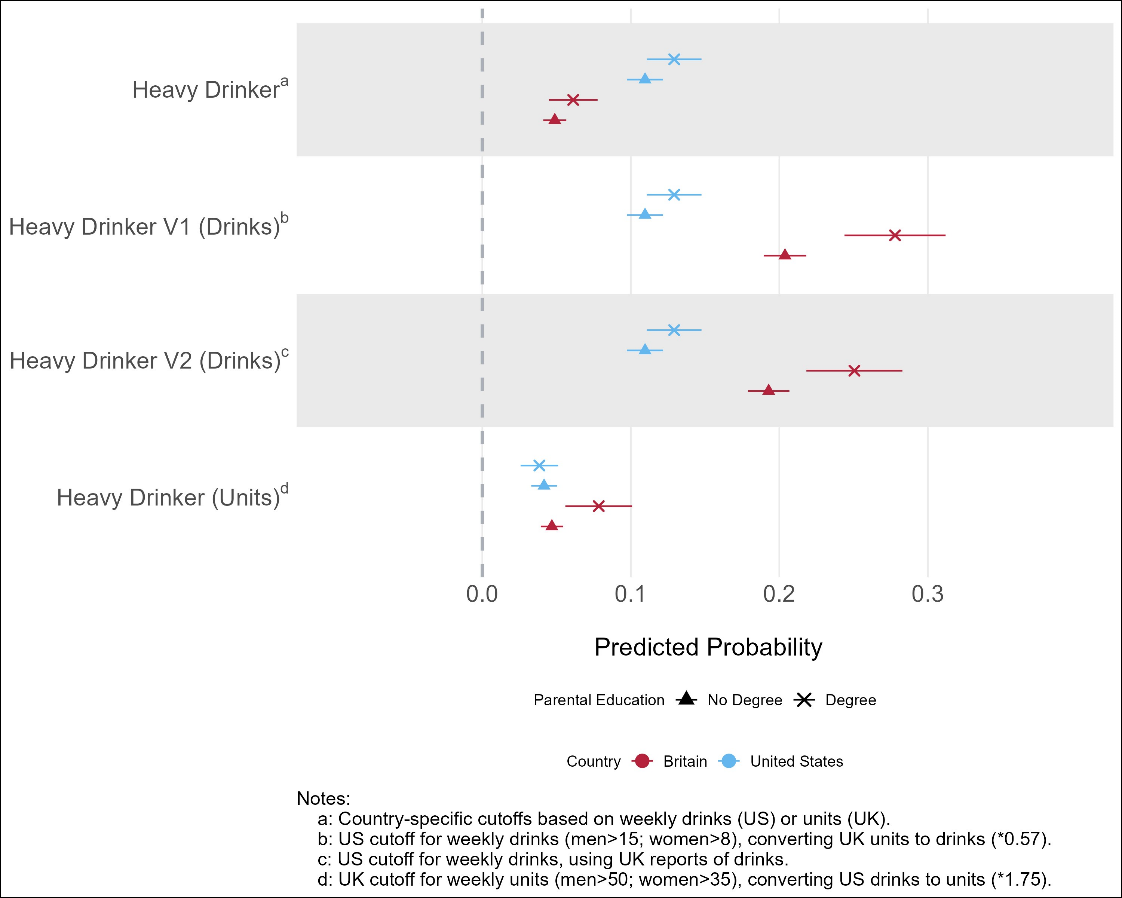

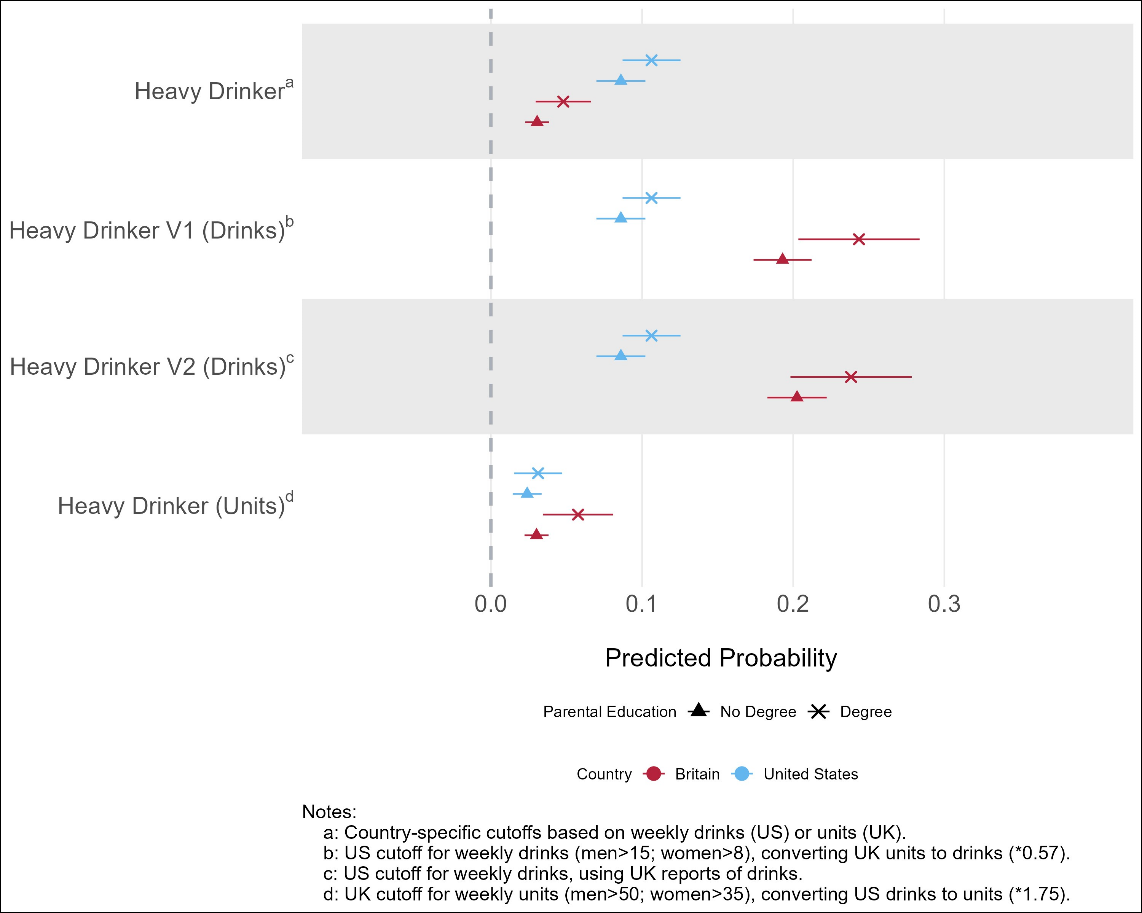
**Supplementary Figure 7f.** Model 3 in females comparing difference operationalisations of heavy drinking.

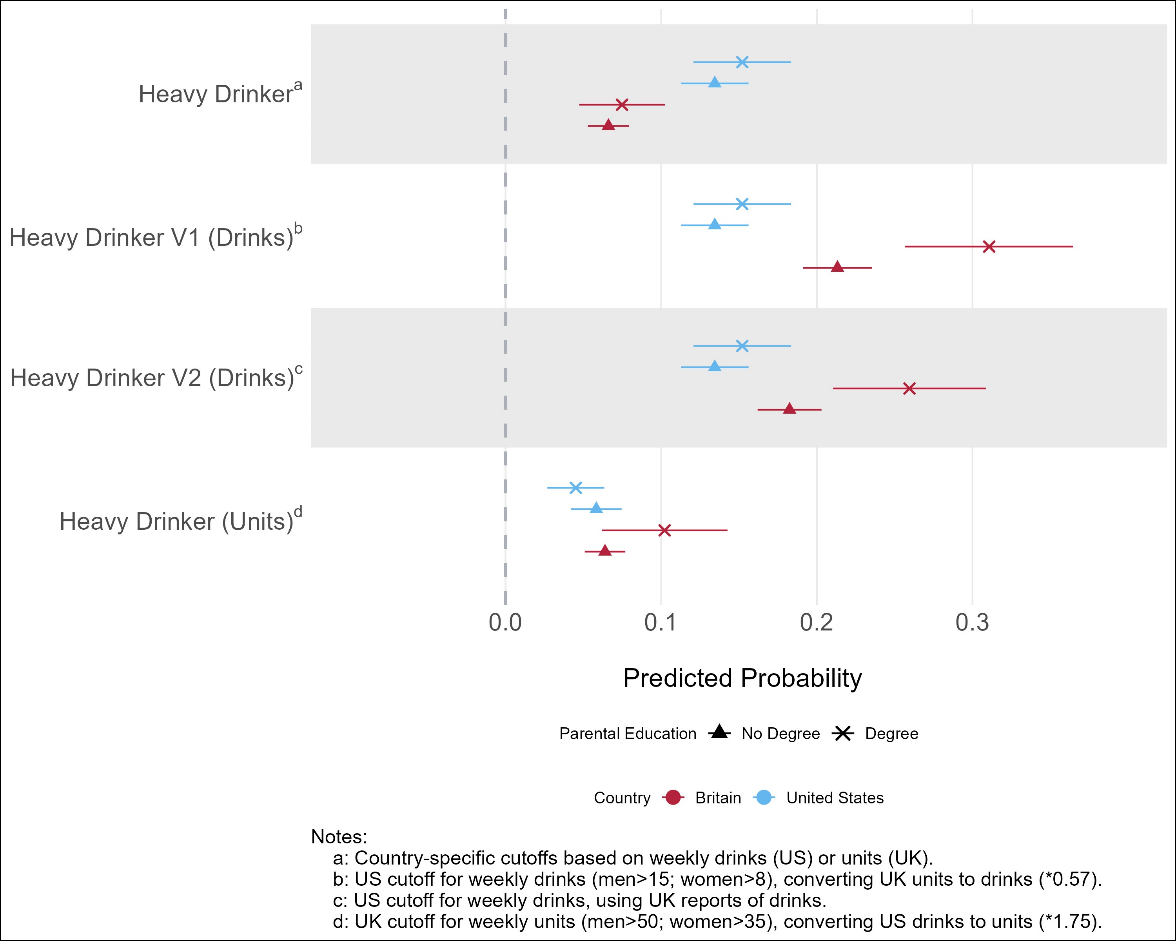
**Supplementary Figure 7g.** Model 3 in males comparing difference operationalisations of heavy drinking.

### **Supplementary Discussion S1: operationalization of heavy drinking.**

In sensitivity analysis (Results S7) we find our results differ based on the operationalization of heavy drinking. Namely, the US guidelines on heavy drinking (15 drinks in men and 8 drinks in women, equivalent to 26 and 14 alcoholic units respectively) are more conservative than in the UK (50 units in men, 35 units in women). The UK definition likely captures a more “extreme” tail of heavy drinking, and there is less evidence of differences between the countries at this end of the distribution. Using the more conservative US definition captures a “lower -level” of heavy drinking, that seems to underlie the higher prevalence seen in Britain when using the US cut-off, particularly among middle- and high-income groups. When adopting country-specific definitions of alcohol consumption, the results show higher levels of heavy drinking among US adults. More broadly, this speaks to the challenge of harmonizing ostensibly straightforward measures like alcohol consumption across countries, given the subjective nature of both interpretation and reporting of relevant survey measures.

**References – Supplementary Material**

1.	Mize TD. Best Practices for Estimating, Interpreting, and Presenting Nonlinear Interaction Effects. Sociol Sci. 2019;6:81-117.

2.	Karaca-Mandic P, Norton EC, Dowd B. Interaction Terms in Nonlinear Models. Health Serv Res. 2012;47(1):255-74.

3.	Long JS, Mustillo SA. Using Predictions and Marginal Effects to Compare Groups in Regression Models for Binary Outcomes. Sociol Method Res. 2021;50(3):1284-320.
